# Genetically predicted levels of the human plasma proteome and risk of stroke: a Mendelian Randomization study

**DOI:** 10.1101/2021.10.22.21265375

**Authors:** Lingyan Chen, James E. Peters, Bram Prins, Elodie Persyn, Matthew Traylor, Praveen Surendran, Savita Karthikeyan, Ekaterina Yonova-Doing, Emanuele Di Angelantonio, David J. Roberts, Nicholas A. Watkins, Willem H. Ouwehand, John Danesh, Cathryn M. Lewis, Paola G. Bronson, Hugh S. Markus, Stephen Burgess, Adam S. Butterworth, Joanna M. M. Howson

## Abstract

Proteins are the effector molecules of biology and are the target of most drugs. To identify proteins and related pathways that may play a causal role in stroke pathogenesis, we used Mendelian randomisation (MR). We tested potential causal effects of 308 plasma proteins (measured in 4,994 blood donors from the INTERVAL study) on stroke outcomes (derived from the MEGASTROKE GWAS) in a two-sample MR framework and assessed whether these associations could be mediated by cardiovascular risk factors. We extended the analysis to identify whether pharmacological targeting of these proteins might have potential adverse side-effects or beneficial effects for other conditions through Phenome-wide MR (Phe-MR) in UK Biobank.

MR showed an association between stroke and genetically predicted plasma levels of TFPI, IL6RA, MMP12, CD40, TMPRSS5 and CD6 (*P*≤1.62×10^−4^). We identified six risk factors (atrial fibrillation, body mass index, smoking, blood pressure, white matter hyperintensities and type 2 diabetes) that were associated with stroke (*P*≤0.0071) using MR. The association of TFPI, IL6RA and TMPRSS5 with stroke could be mediated by these risk factors, such as body mass index, white matter hyperintensity and atrial fibrillation. Thirty-six additional proteins were potentially causal for one or more of these risk factors. The Phe-MR suggested that targeting TFPI could have potential beneficial effects on other disorders of arteries and hyperlipidaemia in addition to stroke. Our results highlight novel causal pathways and potential therapeutic targets for stroke.

## Introduction

Stroke is the second leading cause of death worldwide, estimated to cause ∼5.5 million deaths annually and is the leading cause of long-term disability, with a growing burden on global health ^1^. Therefore, there is a need for new and improved treatments and prevention strategies for stroke. While conventional risk factors, such as hypertension ^2^, account for ∼50% of stroke risk, there remains a need to identify new risk factors, biomarkers and therapies for stroke^3^. In 2017, ∼75% of FDA-approved drugs were targeted at human proteins^4^. Plasma proteins play a central role in a range of biological processes frequently dysregulated in diseases ^5-8^, and represent a major source of therapeutic targets for many indications ^4,9,10^. In particular, plasma proteins are particularly relevant for circulatory diseases such as stroke as they are in physical contact with the blood vessels (compared to tissue-specific diseases, *e*.*g*. inflammatory bowel disease).

Genome-wide association studies (GWAS) of plasma protein levels have identified genetic variants that are associated with proteins, usually referred to as ‘protein quantitative trait loci (pQTLs)’ ^11-13^, offering an opportunity to test the causal effect of potential drug targets on the human disease phenome using Mendelian randomization (MR) ^14,15^. Briefly, MR can be thought of as nature’s randomized trial, by capitalising on the random allocation of genetic variants at conception to separate individuals into subgroups (one equivalent to placebo and the other to intervention in a randomized control trial, RCT) and so allows testing of the potential causal association of risk factors (*e*.*g*. plasma proteins) with disease outcomes (*e*.*g*. stroke) as confounders should also be randomised.

Here, we perform a two-sample MR to estimate the causal effects of plasma proteins on stroke, where we derived genetic instrumental variables of 308 circulating plasma proteins from 4,994 participants ^16^ and obtained genetic associations of stroke subtypes, (any stroke (AS), any ischemic stroke (IS), large-artery-stroke (LAS), cardio-embolic-stroke (CES) and small-vessel-stroke (SVS)) from the MEGASTROKE GWAS ^17^. Then, to verify the robustness of the proteins’ instrumental variables, we perform colocalization analyses. We evaluate the causal relationship of plasma proteins on stroke risk factors and assess potential safety effects of targeting the proteins for stroke therapy by performing a phenome-wide MR in UK Biobank GWASs ^18^.

## Methods

The overall study design is illustrated in **Figure 1**. Details of the methods and study participants are provided below.

**Figure 1.**
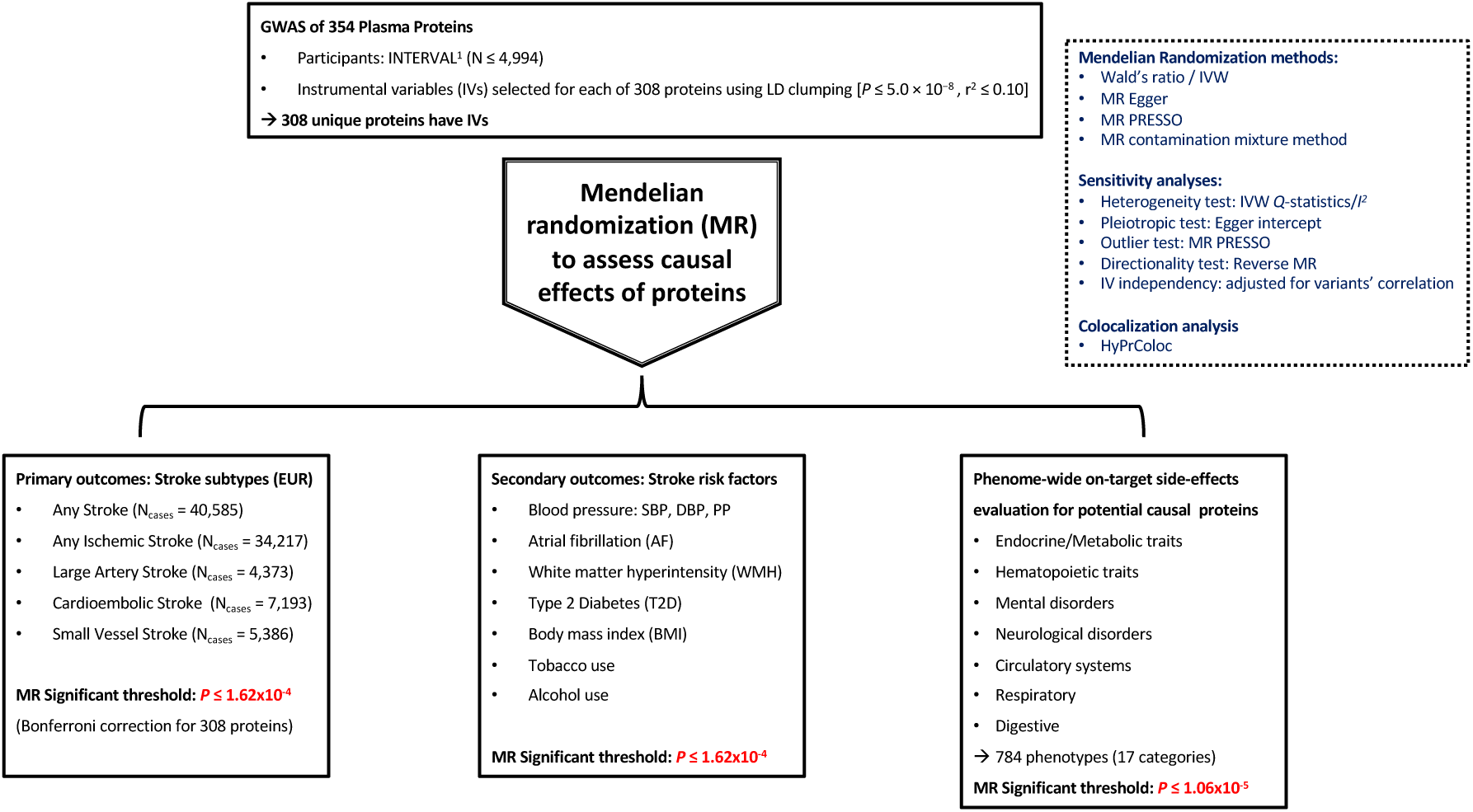
Overview of MR analyses. Four O-link panels were used to measure plasma proteins in a subset of ∼5000 samples from the INTERVAL study ^16^. Genetic variants associated with plasma protein levels were identified based on results from their corresponding GWAS. These genetic variants were then used as proxies for the protein level and tested their relationship with stroke was tested used data from the MEGASTROKE consortium ^17^ for stroke outcomes (Primary MR), with conventional stroke risk factors (Secondary MR), and with 784 phenotypes (Phe-MR) in UK Biobank to test a broad spectrum of potential effects of hypothetical therapeutic agents for stroke.

### Proteomic profiling and quality control

A subset of 4,994 blood donors at mean age of 61 years (SD 6.7 years) enrolled in the INTERVAL BioResource^16^, were processed for proteomic profiling using the Olink Proseek® Multiplex platform by 4 high-throughput, multiplex immunoassays: Inflammatory I (INF1), Cardiovascular II (CVD2), Cardiovascular III (CVD3) and Neurology I (NEURO) (Olink Bioscience, Uppsala, Sweden). Each panel enables the simultaneous measurement of 92 proteins through relative quantification using the Proximity Extension Assay (PEA) Technology ^19^, in which each pair of oligonucleotide-labelled antibodies (“probes”) are allowed to bind to their respective target present in the sample and trigger extension by DNA polymerase. DNA barcodes unique to each protein are then amplified and quantified using a standard real□ time polymerase chain reaction (PCR). Default pre-processing of the proteomic data by Olink included applying median centring normalization between plates, where the median is centred to the overall median for all plates, followed by log2 transformation to provide normalised protein expression (NPX) values. Further details on the Olink proteomic data processing can be found at http://www.olink.com. Probes were labelled using Uniprot identifiers, which we re-mapped to HUGO gene name nomenclature for the (cis-) gene encoding the relevant protein. All protein names and descriptions are provided in **Supplementary Table 1**.

**Table 1.** Data sources for the Mendelian Randomization analysis for current study.

| **Phenotype** | **Source** | **N (Total or Cases/Controls)** | **Imputation reference panel** | **Ancestry** |
| --- | --- | --- | --- | --- |
| **Olink protein levels** | INTERVAL study (unpublished data) | 4,994 | 1000 Genomes Phase 3 + UK10K | European |
| Inflammation Panel (INF1) |  |  |  |  |
| Cardiovascular Panels (CVD2/CVD3) |  |  |  |  |
| Neurology Panel (NEURO) |  |  |  |  |
| **Primary Outcomes** |  |  |  |  |
| Any stroke | 17 studies (Malik et al) <sup>17</sup> | 40,585/406,111 | 1000 Genomes Phase 1 | European |
| Ischemic stroke |  | 34,217/406,111 |  |  |
| Large artery stroke |  | 4,373/406,111 |  |  |
| Cardio-embolic stroke |  | 7,193/406,111 |  |  |
| Small vessel stroke |  | 5,386/406,111 |  |  |
| **Secondary Outcomes** |  |  |  |  |
| Atrial fibrillation (AF) | 6 Studies (Nielsen, et al) <sup>28</sup> | 60,620/970,216 | HRC* | European |
| Type 2 Diabetes (T2D) | 32 Studies (Mahajan, et al) <sup>29</sup> | 74,124/824,006 | HRC | European |
| Body Mass Index (BMI) | GIANT + UK Biobank (Pulit, et al) <sup>31</sup> | 694,649 | HRC | European |
| Tobacco and alcohol use | 29 Studies (Liu, et al) <sup>32</sup> |  | HRC | European |
| AgeSmk |  | 341,427 |  |  |
| CigDay |  | 337,334 |  |  |
| SmkCes |  | 547,219 |  |  |
| SmkInit |  | 1,232,091 |  |  |
| DrnkWk |  | 941,280 |  |  |
| Blood pressure (BP) | UK Biobank (Surendran, et al.) <sup>27</sup> | 445,415 | HRC | European |
| Systolic BP |  |  |  |  |
| Diastolic BP |  |  |  |  |
| Pulse pressure (PP) |  |  |  |  |
| White Matter Hyperintensity (WMH) | UK Biobank + CHARGE + study in stroke patients (Persyn et al., 2020) <sup>30</sup> | 42,310 | HRC | Trans-ethnic, mainly European |
| **On-target side effects evaluation** |  |  |  |  |
| 784 Phenotypes | UK Biobank (Zhou, et al) <sup>33</sup> | 408,961 | HRC | European |

\*HRC: The Haplotype Reference Consortium (HRC); AgeSmk: Age of Initiation of Regular Smoking; CigDay: Cigarettes per day; SmkCes: Smoking Cessation; SmkInit: Smoking Initiation; DrnkWk: Drinks per week.

Samples that failed standard Olink quality control metrics were removed. 4,902, 4,947, 4,987, and 4,660 samples passed quality control for the ‘INF1’, ‘CVD2’, ‘CVD3’ and ‘NEURO’ panels, respectively. According to the manufacturer’s recommendation, we also removed four proteins (HAGH, BDNF, GDNF and CSF3) in the ‘NEURO’ panel and one protein (GDNF) in the ‘INF1’ panel due to high levels of missingness.

### Proteome GWAS

The INTERVAL study^16^ was genotyped using the UK Biobank Affymetrix Axiom array (http://www.ukbiobank.ac.uk/scientists-3/uk-biobank-axiom-array/), and imputed to 1000 Genomes Phase 3-UK10K combined reference panel, employing the PBWT imputation algorithm ^20^. Genetic data for the ∼5000 participants with Olink proteomic profiling were extracted to test for association of the genetic variants with plasma proteins. More details regarding the INTERVAL genetic data QC can be found here ^21^. Within the ∼5,000 participant subset, we removed six related individuals (those individuals with pairwise values of twice the kinship coefficient (PI_HAT) > 0.1875 (removing the individuals with the lowest call rate from each pair). The final imputed dataset was additionally filtered for imputation quality (only retaining variants with an info score > 0.4) and Hardy-Weinberg equilibrium (retaining variants with *P*_HWE_ > 1×10^−4^).

354 proteins (of 363) that passed quality control were taken forward for the GWAS. Normalized protein levels (‘NPX’) were regressed on sex, age, plate, time from blood draw to processing (in days), season (as a categorical variable: ‘Spring’, ‘Summer’, ‘Autumn’, ‘Winter’), and batch when appropriate. The residuals were then rank-inverse normalized. Linear regression of the rank-inversed normalize residuals on genotype was carried out in SNPTEST v.2.5.2 ^22^, with the first three components of multi-dimensional scaling as covariates to adjust for ancestry. Only proteins with at least one SNP with an association *P*-value passing the genome-wide significant threshold (*P* ≤ 5.0×10^−8^) were kept, resulting in 308 proteins for MR analyses.

### Genetic variants associated with proteins

For each plasma protein, cis- and trans-pQTLs from its corresponding GWAS were used as genetic instruments. We followed these steps to select pQTL instruments: (i) we obtained SNPs that were also tested in the MEGASTROKE GWAS of stroke outcomes (see below), (ii) we performed linkage disequilibrium (LD) clumping using PLINK 1.90 (www.cog-genomics.org/plink/1.9/) ^23^ to obtain approximately independent SNPs for each protein. In brief, the LD clumping algorithm groups SNPs in LD (*r*^*2*^ ≥ 0.1 in 4,994 European ancestry participants from the INTERVAL study ^16,21^) within +/-1MB of an index SNP (SNPs with association *P* ≤ 5×10^−8^). Analyses assessing sensitivity to the *r*^*2*^ ≥ 0.1 LD threshold are detailed below. The algorithm loops through all index SNPs, beginning with the smallest *P*-value and only allowing each SNP to appear in one clump. The final output therefore contains the most significant protein-associated SNPs for each LD-based clump across the genome. We split pQTL variants into cis-pQTLs (+/-1MB window of the gene encoding the target protein) and trans-pQTLs (outside the +/-1MB window). We then performed MR in a two-step approach. Our primary analysis was restricted to cis-pQTLs. Having performed MR restricted to cis-pQTL only as IVs, we broadened the analysis to consider the effects of adding in trans-pQTLs as IVs. We estimated the variance of each protein explained by its IVs through calculating the *R*^*2* 24^ and the strength of each IV by the *F-statistic* ^25^. Summary association statistics of all the instrumental variables (IVs) for the 15 significant proteins are provided in **Supplementary Table 2**.

**Table 2.** Proteins representing potential causal factors for stroke and subtypes.

| **Protein** | **N SNPs** | **Outcome** | **OR [95%CI]<sup>#</sup>** | **P value** |
| --- | --- | --- | --- | --- |
| TFPI | 21 | Stroke | 0.949[0.926, 0.972] | $2.01 \times 10^{-5}$ |
| | 21 | Ischemic-stroke | 0.934[0.91, 0.959] | $4.68 \times 10^{-7}$ |
| TMPRSS5 | 20 | Stroke | 1.058[1.039, 1.077] | $1.17 \times 10^{-9}$ |
| | 20 | Ischemic-stroke | 1.059[1.038, 1.08] | $1.36 \times 10^{-8}$ |
| | 20 | Cardioembolic-stroke | 1.089[1.045, 1.134] | $5.33 \times 10^{-5}$ |
| CD40 | 10 | Stroke | 0.919[0.891, 0.949] | $2.05 \times 10^{-7}$ |
| | 10 | Ischemic-stroke | 0.913[0.882, 0.946] | $3.61 \times 10^{-7}$ |
| | 10 | Large-artery-stroke | 0.795[0.723, 0.874] | $2.09 \times 10^{-6}$ |
| CD6 | 23 | Stroke | 1.039[1.021, 1.057] | $1.26 \times 10^{-5}$ |
| IL6RA | 39 | Stroke | 0.972[0.96, 0.984] | $5.43 \times 10^{-6}$ |
| | 39 | Ischemic-stroke | 0.969[0.955, 0.982] | $7.99 \times 10^{-6}$ |
| | 39 | Small-vessel-stroke | 0.939[0.909, 0.97] | $1.60 \times 10^{-4}$ |
| MMP12 | 13 | Stroke | 0.922[0.901, 0.945] | $3.42 \times 10^{-11}$ |
| | 13 | Ischemic-stroke | 0.918[0.895, 0.942] | $9.07 \times 10^{-11}$ |
| | 12 | Large-artery-stroke | 0.793[0.73, 0.861] | $3.53 \times 10^{-8}$ |

<sup>#</sup>OR [95%CI]=Odds ratio and its 95% confidence interval per 1-SD higher genetically-predicted plasma protein level

To assess the robustness of the *r*^*2*^ ≥ 0.1 threshold for IV selection, we performed two additional sensitivity analyses (**Supplementary Table 11**) for proteins of interest to verify the robustness of MR causal relationship: 1) we performed conditional analysis to derive conditionally independent variants as IVs using the FINEMAP software package ^26^ with *--cond* flag; 2) we performed fine-mapping to obtain variants in the 95% credible set as IVs using FINEMAP software package ^26^ with *---sss* flag.

### Genetic variants associated with stroke and its risk factors

The primary outcomes were the risk of stroke and its subtypes. Genetic association estimates for stroke outcomes were obtained from the MEGASTROKE consortium, a large-scale international collaboration launched by the International Stroke Genetics Consortium (ISGC). A detailed description of the study design and characteristics of study participants were provided in the original publication ^17^. To reduce confounding by population stratification, we extracted estimates for the associations of the protein IVs with stroke and its subtypes restricted to individuals of European ancestry (40,585 cases and 406,111 controls). The primary outcomes for this study were any stroke (including both ischemic and haemorrhagic stroke; AS, N_cases_ = 40,585), any ischemic stroke (IS, N_cases_ = 34,217), and the three etiologic ischemic stroke subtypes: large-artery stroke (LAS, N_cases_ = 4,373), cardio-embolic stroke (CES, N_cases_ = 7,193) and small-vessel stroke (SVS, N_cases_ = 5,386). Summary-level data (beta coefficients and standard errors) for the associations of the five stroke outcomes were obtained from the MEGASTROKE GWAS http://www.megastroke.org/index.html.

The secondary outcomes we considered were stroke risk factors, including blood pressure (BP) ^27^, atrial fibrillation (AF) ^28^, type 2 diabetes (T2D) ^29^, white matter hyperintensity (WMH) ^30^, body mass index (BMI) ^31^, alcohol consumption and smoking behaviour ^32^. We used the same pQTLs as IVs for the secondary outcomes as for the primary outcomes. The SNP-outcome effects for all the above risk factors were obtained from previously published GWASs when available. **Table 1** provides full details of the data sources and sample size for these GWASs.

### Phenome-wide MR (Phe-MR) analysis of 784 phenotypes for target proteins

We expanded the exploration of side-effects for the six stroke-associated proteins to include non-stroke phenotypes by performing Phe-MR analyses for a range of diseases. We used summary statistics for SNP-outcome effects calculated using the UK Biobank cohort (N ≤ 408,961) by Zhou et al. ^33^, who performed GWAS using the Scalable and Accurate Implementation of GEneralized mixed model (SAIGE v.0.29) method ^33^ to account for unbalanced case-control ratios. They defined disease outcomes based on “PheCodes”, a system developed to organize International Classification of Diseases and Related Health Problems (ICD-9/-10) codes into phenotypic outcomes suitable for systematic genetic analysis of numerous disease traits ^18,33^. Outcomes with fewer than 500 cases were excluded due to statistical power, leaving 784 diseases for Phe-MR analyses **(Supplementary Table 8**). SNP-outcome associations were downloaded from SAIGE GWAS ^33^ (https://www.leelabsg.org/resources). pQTLs were derived from the same proteome GWAS as in the primary analysis with stroke subtypes. Phe-MR findings can be interpreted as the risk/protective effect per-SD increase in the plasma protein level, same as with primary stroke outcomes. That is, if the effect direction of the additional indication is consistent with the effect direction in Stroke, the identified protein that is therapeutically targeted for the treatment of stroke may also be “beneficial” for the additional indication, and vice versa. MR causal effects are considered statistically significant at *P* ≤ 1.06×10^−5^ (Bonferroni-adjusted for 6 proteins and 784 phenotypes: 0.05/6/784).

### Systematic MR screening for causal proteins of stroke and stroke risk factors

We used two-sample MR ^34-36^ to estimate the associations between genetically-predicted protein levels and target outcomes (stroke, stroke risk factors, and potential adverse effects or additional indications). Two sample MR ^37^ is where the genetic associations with the risk factor are derived in one cohort (*e*.*g*. pQTLs from INTERVAL) and the association of these genetic variants with the outcome is tested in a second cohort (*e*.*g*. stroke GWAS from MEGASTROKE). Two-sample MR allows evaluation of causal effects using summary genetic association data, negating the need for individual participant data. The MR approach was based on the following assumptions: (i) the genetic variants used as instrumental variable (IV) are associated with target exposure, *i*.*e*., protein levels; (ii) there are no unmeasured confounders of the associations between genetic variants and outcome; (iii) the genetic variants are associated with the outcome only through changes in the exposure, *i*.*e*., no pleiotropy.

After extracting the association estimates between the variants and the exposures or the outcomes, we harmonized the direction of estimates by effect alleles, and applied the Wald’s ratio method to estimate the causal effects when there was only one IV available for target exposure. If more than one IV was available, we applied the inverse-variance weighting (IVW) method, either in a fixed-effect framework (IVs ≤ 3) or in a multiplicative random-effect meta-analysis framework (IVs > 3) ^34^. We chose 3 as a cut-off for the random effects model, as with >3 variants, there is potential for some heterogeneity within instrumental variables. (The multiplicative random-effects model allows for heterogeneity between causal estimates targeted by the genetic variants by allowing over-dispersion the regression model.) We also performed several sensitivity analyses to assess the robustness of our results to potential violations of the MR assumptions given these analyses have different assumptions for validity: (i) heterogeneity was estimated by Cochran *Q* test ^34^; (ii) horizontal pleiotropy was estimated using MR-Egger’s intercept ^38^; (iii) influential outlier IVs due to pleiotropy were identified using MR Pleiotropy Residual Sum and Outlier (MR-PRESSO) ^39^; (iv) reverse MR was used to eliminate spurious results due to reverse causation. Additionally, the contamination mixture method ^40^, which can explicitly model multiple potential causal estimates and therefore infer multiple causal mechanisms associated with the same risk factor that affect the outcome to different degrees, was also used to calculate the MR estimates. Although these methods may have differing assumptions and statistical power, the rationale for using them is that if they give a similar conclusion, this provides greater certainty in inferring that any positive results are unlikely to be driven by violation of the MR assumptions.

Effects on binary outcomes (*i*.*e*., stroke, AF, T2D, smoking initiation/cessation) are reported as odds ratios (ORs) with their 95% confidence intervals (CIs) scaled to a one standard deviation (SD) higher plasma protein level. Effects on quantitative outcomes (*i*.*e*., BP, WMH, BMI) are reported as the effect size (95% CI) scaled to a 1-SD higher plasma protein levels. All statistical tests were two-sided and considered statistically significant at *P*_*CausalEstimate*_ ≤ 1.62×10^−4^ (Bonferroni-adjusted for 308 proteins: 0.05/308=1.62×10^−4^), *P*_*Q-stat*_ ≥ 0.05, *P*_*Egger-Intercept*_ ≥ 0.05 and *P*_*GlobalTest*_ ≥ 0.05. The MR analyses were conducted using *MendelianRandomization* (Version: 0.4.2) ^35^, *TwoSampleMR* (Version: 0.4.22) ^36^ and *MR-PRESSO* (Version: 1.0) ^39^ packages in R 3.5.1 (R Foundation, www.R-project.org). Plots were generated using various R packages including *ggplot2* (Version: 3.2.0), *forestplot* (Version: 1.9), and *PheWAS* (Version: 0.99.5-4). We employed the same statistical analysis framework, incorporating the sensitivity analyses for all MR analyses.

### Multi-trait colocalization analyses

As the instruments used in the current setting were identified based on their statistical associations with the protein level, we conducted another sensitivity analysis – colocalization, to investigate whether the genetic associations with both protein and phenotypes shared the same causal variants. We conducted colocalization analysis for each potential causal protein across multiple traits, including protein level and five stroke outcomes, to estimate the posterior probability (PP) of multiple traits sharing the same causal SNP simultaneously using a multi-trait colocalization (HyPrColoc) method ^41^. HyPrColoc extends the established coloc methodology ^42^ by approximating the true posterior probability of colocalization with the posterior probability of colocalization at a single causal variant and a small number of related hypotheses. If all traits do not share a causal variant, HyPrColoc employs a novel branch-and-bound selection algorithm to identify subsets of traits that colocalize at distinct causal variants at the locus. We used uniform priors for the primary analysis. We also performed sensitivity analysis with non-uniform priors to access the choice of priors, which used a conservative trait-level prior structure with *P*=1×10^−4^ (prior probability of a SNP being associated with one trait) and γ=0.98 (1-prior probability of a SNP being associated with an additional trait given that the SNP is associated with at least one other trait), *i*.*e*., 1 in 500,000 variants are expected to be causal for two traits.

Variants within a ±1Mb window around the pQTL with the smallest *P*-value, with imputation (INFO)-score ≥ 0.8 and minor allele frequency (MAF) ≥ 0.01 were included. All variants across each of the datasets were harmonized to the same effect alleles prior to colocalization analyses. We conducted the colocalization analysis using the ‘HyPrColoc’ R package ^41^.

## Results

### Genetically determined plasma protein levels and risk of stroke

Three hundred and eight plasma proteins were tested for causal associations with stroke outcomes (**Figure 1**). As cis-pQTLs were considered to have a more direct and specific biological effect upon the protein (compared to trans-pQTLs) ^43^, we first performed MR analyses using only cis-pQTLs as instrumental variables and identified six putatively causal proteins with at least one stroke outcome (*P* ≤ 1.62×10^−4^ =0.05/308 proteins; **Table 2, Figure 2 & Figure 3, Supplementary Figure 1**): TFPI (Tissue Factor Pathway Inhibitor), TMPRSS5 (Transmembrane Serine Protease 5), CD40 (B Cell Surface Antigen CD40), MMP12 (Matrix Metallopeptidase 12), IL6RA (Interleukin 6 Receptor), and CD6 (T-Cell Differentiation Antigen CD6). TFPI, CD40, IL6RA, and MMP12 were significantly associated with lower risk of any stroke and any ischemic stroke, while TMPRSS5 and CD6 was significantly associated with higher risk of any stroke. Among the ischemic stroke subtypes, genetic predisposition to upregulated TMPRSS5 was associated with higher risk of any ischemic stroke (OR per-1-SD higher plasma protein level [95%CI]=1.059[1.038, 1.08]; *P*=1.36×10^−8^) and Cardioembolic stroke (OR[95%CI]=1.089[1.045, 1.134]; *P*=5.33×10^−5^). Higher genetically predicted levels of both MMP12 (OR[95%CI]=0.793[0.73, 0.861]; *P*=3.53×10^−8^) and CD40 (OR[95% CI]= 0.795[0.723, 0.874]; *P*=2.09×10^−6^) were associated with lower risk of Large-artery stroke. Higher genetically predicted soluble IL6RA (and lower IL6R signalling ^44^) was associated with lower risk of Small-vessel stroke (OR[95% CI]= 0.939[0.909, 0.970]; *P=*1.60×10^−4^).

**Figure 2.**
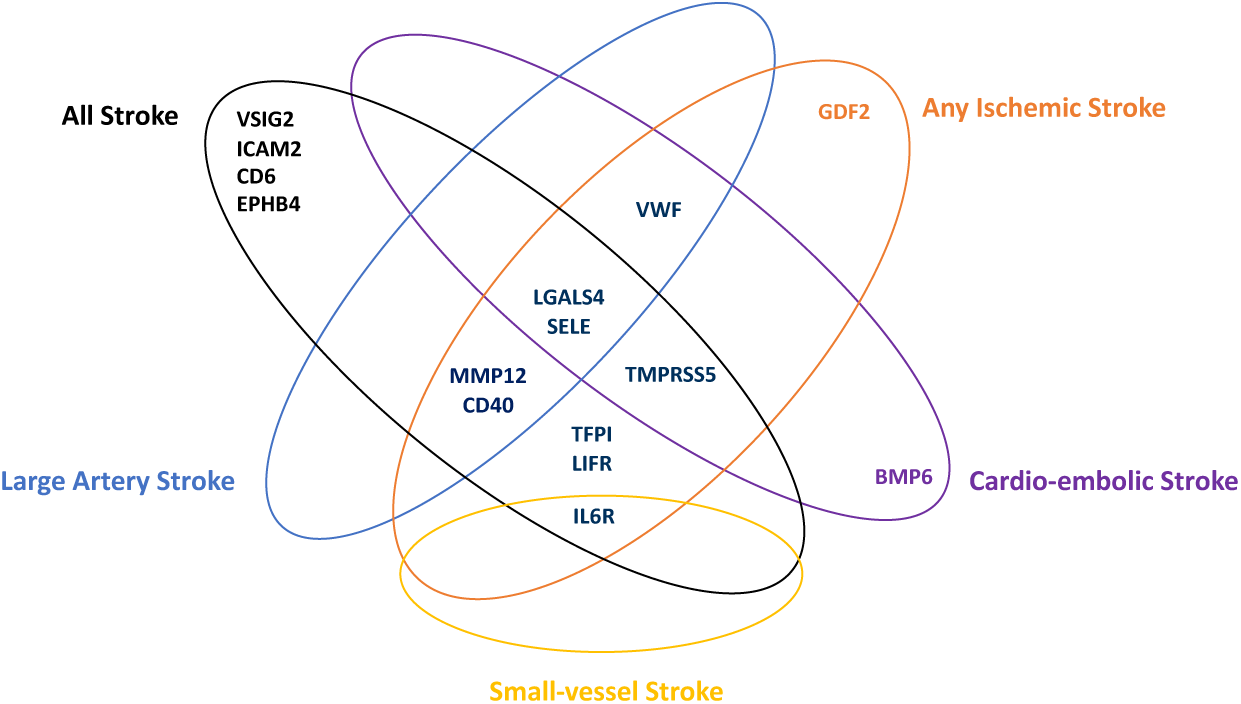
Venn diagram of identified potential causal proteins for stroke subtypes.

**Figure 3.**
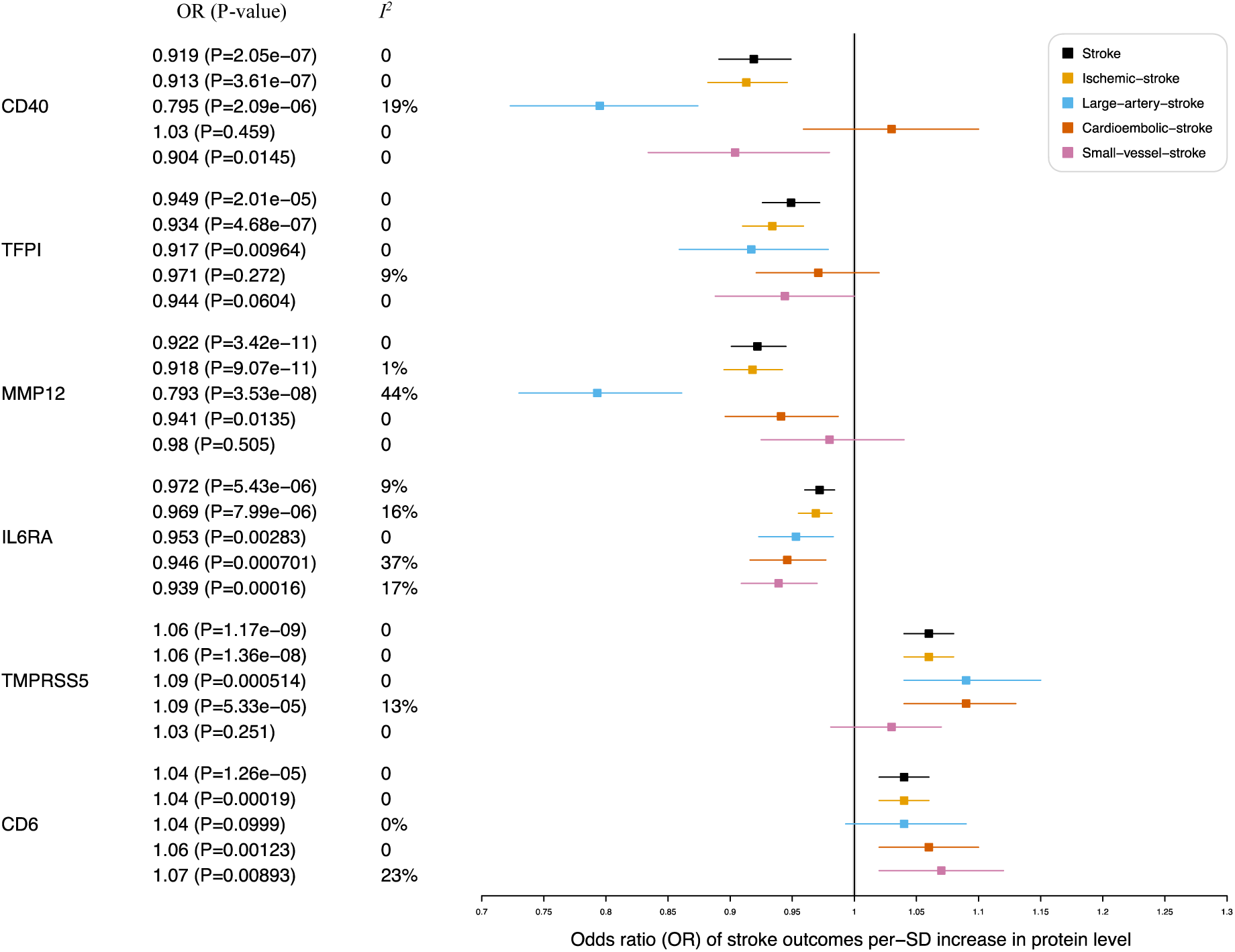
Effects of six potential causal proteins on stroke subtypes. OR: Odds ratio; *I*^*2*^: heterogeneity. CD40: B Cell Surface Antigen CD40; TFPI: Tissue Factor Pathway Inhibitor; MMP12: Matrix Metallopeptidase 12; IL6RA: Interleukin 6 Receptor Subunit Alpha; TMPRSS5: Transmembrane Serine Protease 5; CD6: T-Cell Differentiation Antigen CD6.

We extended the MR analyses to include trans-pQTLs as instrumental variables and identified nine additional proteins significantly associated with at least one stroke outcome (*P* ≤ 1.62×10^−4^; Supplementary **Table 3**). However, seven proteins (VSIG2, EPHB4, Gal4, ICAM2, LIFR, SELE, and vWF), included instrumental variables from the *ABO* locus, which is well known to have pleiotropic effects. We note that the ABO protein has previously been identified as a genetic risk factor for stroke^45^ . Interestingly, both Bone Morphogenetic Protein 6 (BMP6) and Growth Differentiation Factor 2 (GDF2, also known as BMP9) were instrumented by *trans*-pQTLs located in the genetic regions of *KNG1* (Kininogen 1) and *F11* (Coagulation Factor XI). Both genes are essential for blood coagulation and the latter has previously been reported to be a causal risk factor for stroke ^46^. GDF2 has also been found to have a causal role in pulmonary artery hypertension (PAH)^47^. We therefore focused further analyses on the proteins with *cis* pQTL only (*i*.*e*., TFPI, TMPRSS5, CD40, MMP12, IL6RA, CD6), as these associations with stroke are unlikely to be due to pleiotropy.

**Table 3.**

a. MR results of risk factors and stroke outcomes.
| Risk Factors | Stroke outcome | N SNPs | OR [95%CI] * | P-value |
| --- | --- | --- | --- | --- |
| AF | Stroke | 130 | 1.21[1.18, 1.24] | $2.23 \times 10^{-39}$ |
| | Ischemic-stroke | 130 | 1.22[1.19, 1.26] | $1.11 \times 10^{-39}$ |
|  | Large-artery-stroke | 131 | 1.04[0.97, 1.12] | 0.315 |
| | Cardioembolic-stroke | 133 | 2.04[1.92, 2.16] | $2.72 \times 10^{-125}$ |
|  | Small-vessel-stroke | 131 | 0.99[0.92, 1.06] | 0.714 |
| BMI | Stroke | 792 | 1.19[1.13, 1.25] | $1.73 \times 10^{-10}$ |
| | Ischemic-stroke | 791 | 1.17[1.11, 1.24] | $3.25 \times 10^{-08}$ |
| | Large-artery-stroke | 794 | 1.29[1.12, 1.49] | $3.24 \times 10^{-04}$ |
| | Cardioembolic-stroke | 792 | 1.15[1.04, 1.28] | $7.49 \times 10^{-03}$ |
|  | Small-vessel-stroke | 794 | 1.11[0.98, 1.25] | 0.118 |
| SBP | Stroke | 701 | 1.67[1.57, 1.78] | $1.37 \times 10^{-56}$ |
| | Ischemic-stroke | 704 | 1.68[1.57, 1.81] | $3.49 \times 10^{-48}$ |
| | Large-artery-stroke | 710 | 2.58[2.21, 3.01] | $3.69 \times 10^{-33}$ |
| | Cardioembolic-stroke | 705 | 1.36[1.2, 1.53] | $8.31 \times 10^{-07}$ |
| | Small-vessel-stroke | 708 | 1.99[1.72, 2.29] | $3.66 \times 10^{-21}$ |
| DBP | Stroke | 661 | 1.5[1.4, 1.6] | $1.04 \times 10^{-32}$ |
| | Ischemic-stroke | 667 | 1.5[1.4, 1.62] | $1.67 \times 10^{-28}$ |
| | Large-artery-stroke | 674 | 1.72[1.46, 2.02] | $5.01 \times 10^{-11}$ |
| | Cardioembolic-stroke | 673 | 1.26[1.12, 1.43] | $2.11 \times 10^{-04}$ |
| | Small-vessel-stroke | 677 | 1.8[1.54, 2.1] | $6.30 \times 10^{-14}$ |
| PP | Stroke | 723 | 1.4[1.31, 1.49] | $1.07 \times 10^{-25}$ |
| | Ischemic-stroke | 723 | 1.41[1.33, 1.51] | $1.82 \times 10^{-25}$ |
| | Large-artery-stroke | 725 | 2.23[1.92, 2.59] | $3.29 \times 10^{-26}$ |
| | Cardioembolic-stroke | 726 | 1.19[1.06, 1.33] | $3.21 \times 10^{-03}$ |
| | Small-vessel-stroke | 725 | 1.53[1.34, 1.75] | $4.17 \times 10^{-10}$ |
| T2D | Stroke | 340 | 1.09[1.07, 1.12] | $2.23 \times 10^{-13}$ |
| | Ischemic-stroke | 341 | 1.1[1.08, 1.13] | $3.96 \times 10^{-15}$ |
| | Large-artery-stroke | 341 | 1.24[1.16, 1.32] | $1.76 \times 10^{-11}$ |
|  | Cardioembolic-stroke | 337 | 1.05[1, 1.1] | 0.0352 |
| | Small-vessel-stroke | 342 | 1.17[1.1, 1.24] | $1.05 \times 10^{-07}$ |
| WMH | Stroke | 16 | 1.21[1.07, 1.36] | $2.21 \times 10^{-03}$ |
|  | Ischemic-stroke | 17 | 1.15[1, 1.31] | 0.046 |
|  | Large-artery-stroke | 18 | 1.13[0.9, 1.43] | 0.295 |
|  | Cardioembolic-stroke | 18 | 1.14[0.94, 1.37] | 0.183 |
| | Small-vessel-stroke | 15 | 1.49[1.17, 1.9] | $1.47 \times 10^{-03}$ |
| SmkInit | Stroke | 365 | 1.24[1.16, 1.32] | $1.80 \times 10^{-11}$ |
| | Ischemic-stroke | 364 | 1.22[1.15, 1.31] | $6.34 \times 10^{-10}$ |
| | Large-artery-stroke | 365 | 1.66[1.42, 1.93] | $8.88 \times 10^{-11}$ |
|  | Cardioembolic-stroke | 365 | 1.11[0.98, 1.25] | 0.116 |
| | Small-vessel-stroke | 365 | 1.46[1.25, 1.71] | $1.27 \times 10^{-06}$ |

**b. MR results of stroke associated proteins and risk factors.**
| Protein | N SNPs | Risk Factors | OR/$\beta$ [95%CI] <sup>#</sup> | P value |
| --- | --- | --- | --- | --- |
| TFPI | 19 | BMI | -0.013[-0.019, -0.007] | $3.56 \times 10^{-5}$ |
| | 19 | WMH | -0.06[-0.08, -0.04] | $7.15 \times 10^{-10}$ |
| TMPRSS5 | 21 | AF | 1.03[1.016, 1.045] | $2.15 \times 10^{-5}$ |
| IL6RA | 46 | AF | 0.959[0.95, 0.968] | $2.55 \times 10^{-18}$ |

<sup>#</sup>OR/ $\beta$ [95%CI]: Odds ratio/log(odds ratio) and its 95% confidence interval, indicates pe-1-SD increase in protein level and its effect on each outcome, if the outcome is a continuous trait, *e.g.* BMI, BP, we report the effect using $\beta$ , otherwise, we use OR (for binary outcome). OR=Odds Ratio; CI=Confidence Interval; AF=Atrial Fibrillation; BMI=Body Mass Index; WMH=White Matter Hyperintensity; T2D=Type 2 Diabetes; SBP=Systolic Blood Pressure; DBP=Diastolic Blood Pressure; PP=Pulse Pressure; SmkInit=Smoking Initiation.

Results of sensitivity analyses confirmed the robustness of the primary MR analyses. There was no evidence for heterogeneity in the association of any of the six proteins in **Supplementary Table 3** as measured by Cochran Q statistics (*P*_*Q-stat*_ > 0.05), and no indication that the instrumental variables had horizontal pleiotropy as assessed by MR-Egger intercept (*P*_*Egger-Intercept*_ > 0.05) or MR-PRESSO global pleiotropy test (*P*_*GlobalTest*_ > 0.05). All MR causal effect estimates adjusting for correlation of IVs’ were consistent with the primary analyses (**Supplementary Table 10**). Moreover, MR causal estimates using IVs derived from conditionally independent variants and credible sets of variants from fine-mapping showed consistent results (**Supplementary Table 11 & 12**). There was no evidence of reverse causations (**Supplementary Table 13**).

### Co-localization

We formally tested whether the associations of the variant with the protein levels used as IVs and the stroke outcome are shared for the six proteins using statistical colocalization analysis. We applied a Bayesian algorithm, Hypothesis Prioritization in multi-trait Colocalization (HyPrColoc), which allows for the assessment of colocalization across multiple complex traits simultaneously (**Methods**), to test whether the protein associations and stroke associations are shared. The association of the genetic variants selected as instrumental variables for four proteins (TFPI, TMPRSS5, CD40, and CD6) colocalized with the stroke associations (posterior probability (PP) ≥ 0.7) (**Supplementary Table 4, Supplementary Figure 2)** *i*.*e*., the associations in these regions were likely due to the same underlying causal variants. The colocalization suggested the genetic variants associated with TFPI (pQTLs) were due to the same genetic variants underlying the association with all-stroke. Similarly, CD6 pQTLs colocalized with all-stroke genetic associations; CD40 pQTLs colocalized with the genetic associations for all-stroke, ischemic-stroke and large-artery-stroke; TMPRSS5 pQTLs colocalized with all-stroke, ischemic-stroke and cardioembolic-stroke genetic associations. Notably, we found for TFPI, CD40, and CD6 that >80% of the posterior probability of colocalization of the primary genetic association with stroke and the respective protein levels were explained by a single variant (rs67492154, rs4810485, and rs2074227 for TFPI, CD40, and CD6, respectively). The colocalization evidence at MMP12, was less strong than with the other proteins, with colocalization PP>0.6 and there was no colocalization evidence for IL6RA with stroke, which could be due to violation of the single causal variant assumption of the HyprColoc method.

### Characterizing the potential causal effects of stroke risk factors on stroke

To understand potential causal mechanisms between plasma proteins and stroke, we conducted mediation MR analyses for conventional stroke risk factors. First, we performed two-sample MR analyses to characterize the causal relationship of the stroke risk factors with all stroke outcomes. Second, we assessed the causal effects of the proteins on the highlighted risk factors.

For each of the six stroke risk factors we considered (*i*.*e*., blood pressure (BP), atrial fibrillation (AF), type 2 diabetes (T2D), white matter hyper-intensity (WMH), body mass index (BMI), smoking behaviours and alcohol consumption), instrumental variables were derived from published GWAS summary statistics restricted to European populations (**Table 1 & Supplementary Table 5**). AF, T2D, smoking, increased systolic BP, diastolic BP, pulse pressure, WMH, and BMI significantly increased the risk of any stroke (*P*≤0.05/7=0.007, Bonferroni adjusted for seven risk factors; **Figure 4, Supplementary Table 6 & Supplementary Figure 3**). As expected, systolic BP exhibited the strongest effect of all the risk factors on any ischemic stroke and LAS (OR per-1-SD [95% CI]=1.68[1.57, 1.80] and 2.58[2.21, 3.01], respectively) and AF had a positive association with CES (OR[95% CI]: 2.04[1.92, 2.16]; *P*=2.72×10^−125^). WMH increased risk of any stroke and SVS (1-SD increased in WMH was associated with 49% higher odds for SVS (OR[95% CI]=1.49[1.17, 1.9]; *P*=0.00147). Both genetically determined higher T2D risk and smoking initiation were associated with increased risk of LAS and SVS; and genetically determined higher BMI was associated with higher risk of LAS. No significant association was observed for alcohol consumption with any of the stroke outcomes (*P*>0.05).

**Figure 4.**
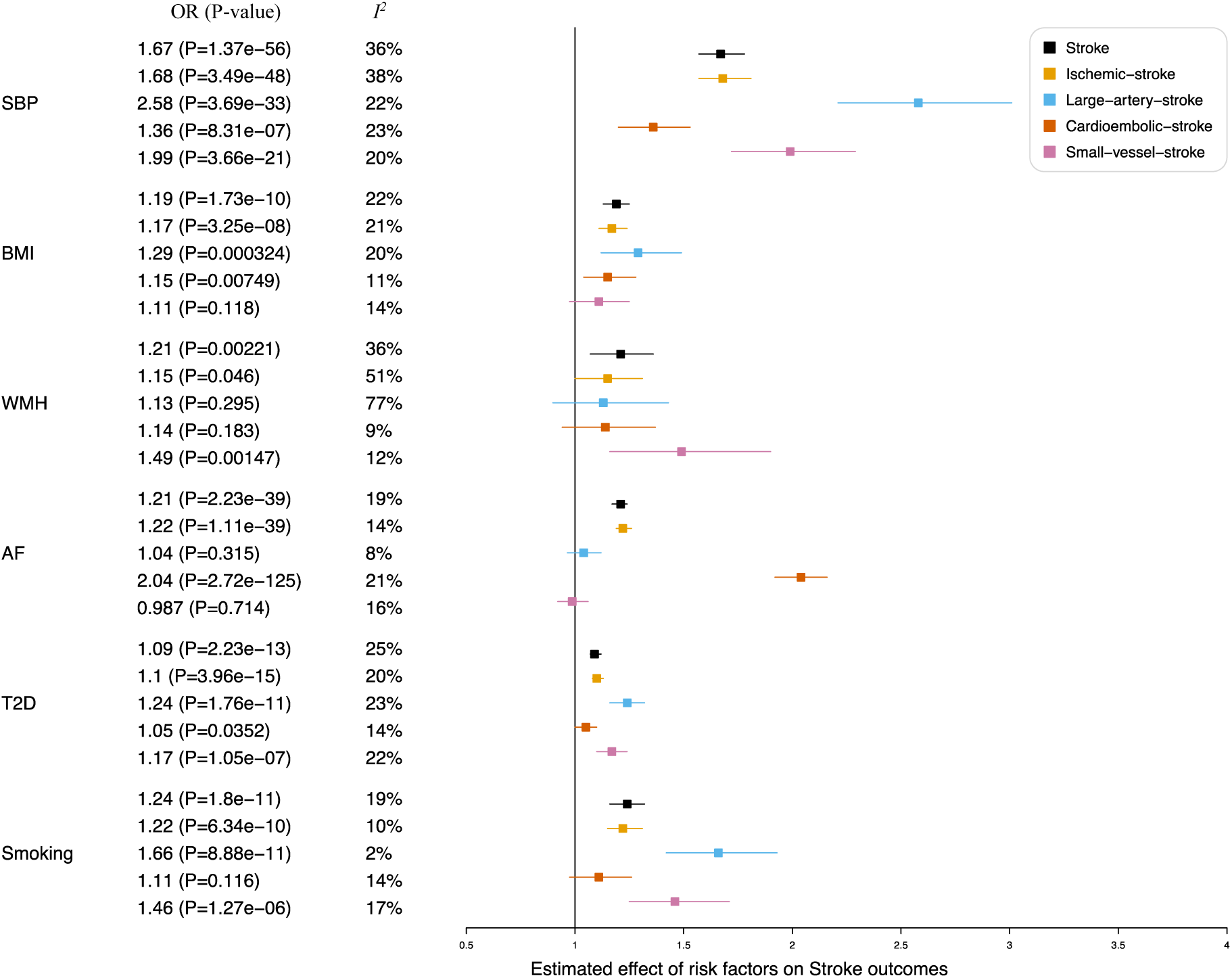
Causal effects of risk factors on stroke subtypes. *I*^*2*^: heterogeneity. SBP = Systolic Blood Pressure; AF = Atrial Fibrillation; WMH = White Matter Hyperintensity; T2D = Type 2 Diabetes; BMI = Body Mass Index; Smoking = Smoking Initiation.

### Associations of genetically determined plasma protein levels with stroke risk factors

We performed MR of all 308 plasma proteins with the highlighted stroke risk factors (excluding alcohol consumption which was not associated with increased stroke risk in the above MR analyses). After multiple testing correction, 39 proteins instrumented with cis-pQTLs were significantly associated with at least one stroke risk factor (*P* ≤ 1.62×10^−4^): 5 with Systolic BP; 7 with Diastolic BP; 7 with Pulse Pressure; 6 with AF; 4 with T2D; 9 with BMI; 3 with WMH; and 8 with smoking. There was no evidence of horizontal pleiotropy, and sensitivity analyses yielded consistent causal effect estimates (**Supplementary Table 14**).

Among the six stroke-associated proteins, three proteins were found to be significantly associated with one or more of the stroke risk factors (**Figure 5**; **Table 3**; **Supplementary Figure 4**). Of note, we found genetically predicted higher TFPI level was associated with lower WMH and lower BMI (a 0.06 SD lower WMH β[95% CI]= -0.06[-0.08, -0.04]; *P*=7.15×10^−10^ and a 0.013 SD lower BMI β[95% CI]= -0.013[-0.019, -0.007]; *P*=3.56×10^−5^ per-SD higher TFPI; **Supplementary Table 7**). We thus inferred that the association between TFPI and stroke could be partially mediated by BMI and WMH. Genetically determined higher TMPRSS5 levels were associated with higher risk of AF (OR[95% CI]: 1.03[1.016, 1.045]; *P*=2.15×10^−5^). Together with the causal relationship of AF and cardioembolic stroke, we can also infer that AF is a possible mediator on the effect of TMPRSS5 on cardioembolic stroke. Genetically higher IL6RA levels were associated with a 4.1% lower risk of AF (OR[95% CI]: 0.96[0.95, 0.97]; *P*=2.55×10^−18^). All the effect directions of these associations of proteins with risk factors were consistent with those of the proteins with stroke, indicating that these risk factors may be potential mediators of the protein-stroke associations.

**Figure 5.**
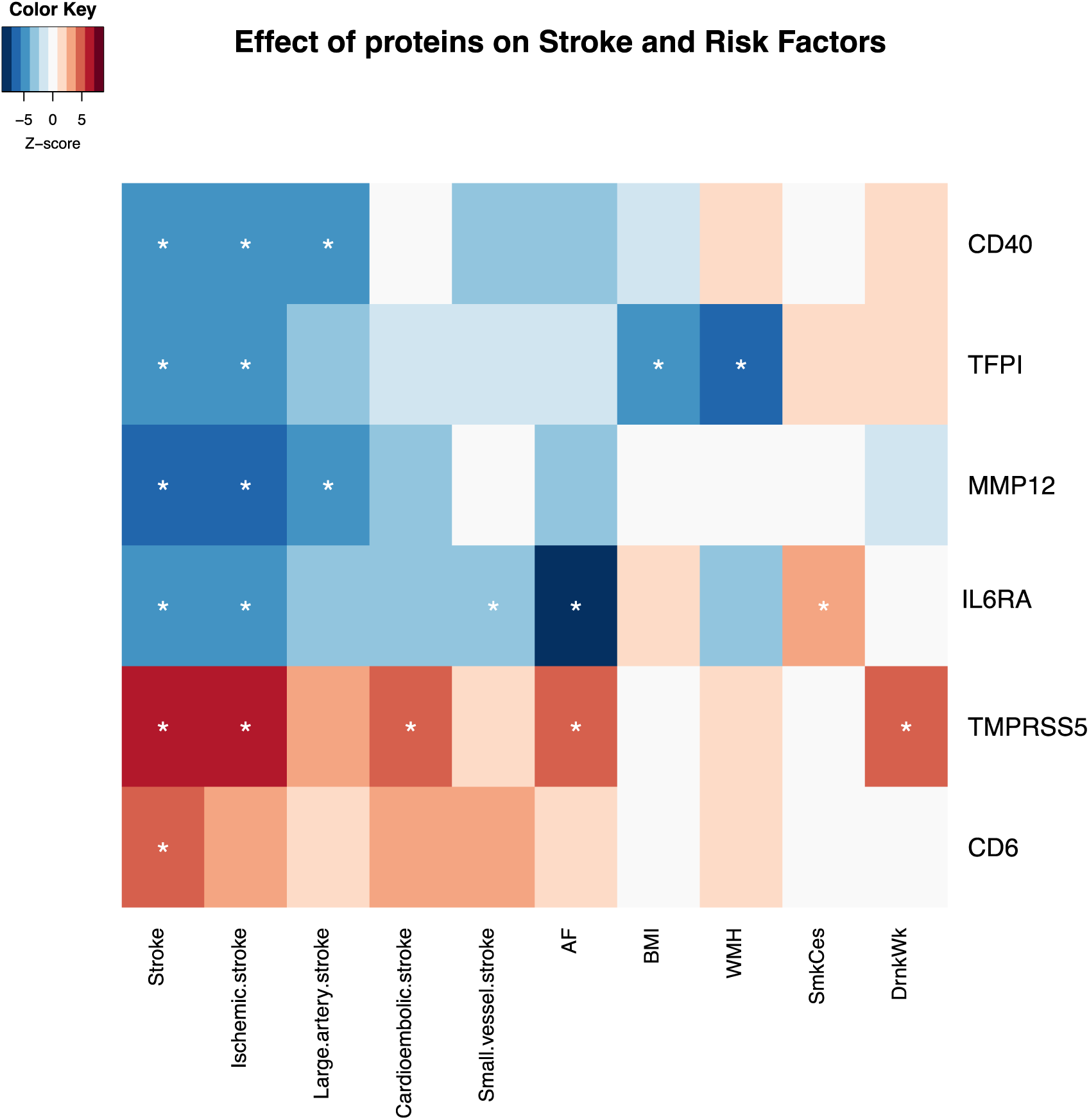
Effect sizes (Z-score) of six potential causal proteins on stroke subtypes and causal risk factors for stroke. Colours in each lattice of the heatmap represent the effect size (Z-score), with genetically predicted increased protein level associated with higher risk of outcomes coloured in brown and lower risk of outcomes coloured in blue. The darker the colour the larger the effect size. * indicates that the causal association is significant, which passed Bonferroni correction of *P*_*causalEstimate_IVW*_ ≤ 0.05/308 =1.61×0^-4^ and passed sensitivity tests with *P*_*Qstat*_ ≥ 0.05 and *P*_*EggerIntercept*_ ≥ 0.05.

Among the 39 proteins that were associated with at least one stroke risk factor, 36 were found to be associated with the risk factors but not stroke outcome (**Supplementary Table 14**). For example, genetically determined Fibroblast Growth Factor 5 (FGF5) level was associated with higher risk of AF (OR=1.056 per SD higher FGF5); each SD higher genetically determined Glypican 5 (GPC5) was associated with higher risk of T2D (OR=1.02); each SD higher in genetically determined Scavenger Receptor Class F Member 2 (SCARF2) was associated with a 0.062-SD higher WMH. We found that higher genetically determined Alpha-L-Iduronidase (IDUA) and Sialic Acid-Binding Ig-Like Lectin 9 (SIGLEC9) were both associated with lower BMI. Higher genetically determined Serine Protease 27 (PRSS27) was associated with higher SBP, higher DBP and higher PP, while higher genetically determined levels of Neurocan (NCAN) were associated with lower risk of T2D (OR=0.76) and 0.07-SD lower SBP.

### Phenome-wide MR (Phe-MR) analysis of stroke-associated proteins in UK Biobank

To assess whether the six stroke-associated proteins have either beneficial or deleterious effects for other indications, we performed a broader MR screen of 784 diseases and traits in UK Biobank (**Supplementary Table 8**). Our Phe-MR results can be interpreted as a per-SD increase in genetically determined plasma protein level that leads to an higher or lower odds of a given disease or trait. If the effect direction of the protein on the disease or trait is the same as on stroke, the effect is considered “beneficial” and “deleterious” otherwise. Overall, 34 significant associations were identified (*P* ≤ 0.05/6/784 =1.06×10^−5^), of which 21 (61.7%) were in the same direction as the stroke association **(Supplementary Table 9**).

Notably, genetically higher levels of plasma TFPI were not only associated with lower risk of stroke, but also lower risk of other diseases involving the circulatory system (cerebrovascular disease, other disorders of arteries), metabolic traits (hyperlipidemia and hypercholesterolemia, disorders of lipid metabolism) and digestive system disorders (acute gastritis); however, they were also associated with higher risk of excessive or frequent menstruation (**Figure 6 & Supplementary Figure 5**). Genetically higher levels of plasma TMPRSS5 were associated with higher risk of cardioembolic stroke, as well as protein−calorie malnutrition (metabolic trait) (**Figure 6 & Supplementary Figure 5**). All the significant associations for CD40, including haemoptysis and abnormal sputum (respiratory system) were consistent with the effect direction of that with stroke. Effects of IL6RA on risk of diseases on circulatory system disorders (ischemic heart disease, cardiac dysrhythmias, atrial fibrillation and flutter, coronary atherosclerosis, angina pectoris, abdominal aortic aneurysm) and musculoskeletal disease (other inflammatory spondylopathies) were consistent with that on risk of stroke; but had inverse effects on dermatologic symptoms (*e*.*g*. cellulitis and abscess of arm/foot), digestive system (*e*.*g*. cholelithiasis) and chronic renal failure [CKD] (**Supplementary Figure 6 & Supplementary Table 9**). Genetically predicted CD6 was associated with alcoholic liver damage and degeneration of intervertebral disc (musculoskeletal system) but in the inverse direction to stroke. Summary results of the primary and sensitivity MR analyses for all the 784 phenotypes are provided in **Supplementary Table 9**.

**Figure 6.**
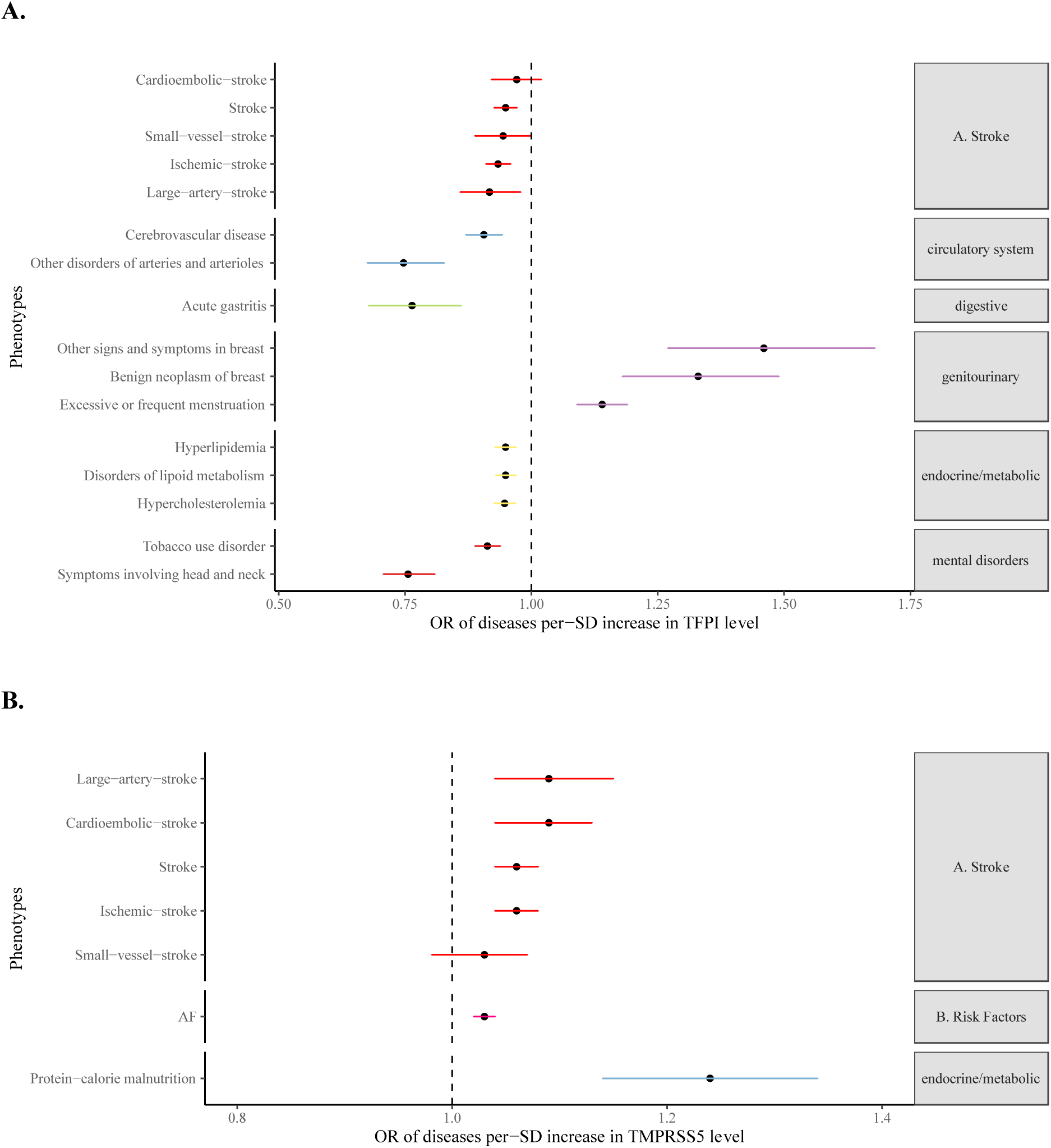
Forest plots illustrating the potential on-target side-effects associated with causal proteins revealed by Phe-MR analysis for TFPI (A) and TMPRSS5 (B). In general, results can be perceived as the effects of per SD higher circulating protein level on each phenotype. If the effect direction of the target protein on the phenotype is consistent with that on stroke outcomes, it represents “beneficial” additional indications through intervention of circulating protein level. Conversely, opposing effect directions of the target protein on the phenotype and stroke represents “deleterious” side-effects. For example, a higher level of TFPI is associated with lower risk of ischemic stroke and so phenotypes with OR<1 represents “beneficial effects”, OR>1 represents “deleterious effects” when the hypothetical intervention increases TFPI levels. Only significant associations that passed Bonferroni correction (*P* ≤ 0.05/6/784 =1.06×10^−5^) were plotted. See **Supplementary Table 9** for more clinical information of the ICD code phenotypes.

## Discussion

Based on genetic data for 308 proteins involved in cardiovascular disease, inflammation and neurological processes from ∼5000 individuals ^16^, our study provides robust evidence that six proteins (TFPI, TMPRSS5, CD40, MMP12, IL6RA, and CD6) are causally associated with stroke. We showed that AF, systolic and diastolic BP, BMI, T2D, WMH and smoking were causally associated with risk of any stroke (and some ischemic stroke subtypes), demonstrating a key role of the risk factors in the pathogenesis of stroke consistent with classical epidemiological data ^48-56^. We found the associations of TFPI, IL6RA, and TMPRSS5 with stroke were likely to be mediated by one or more of these risk factors. In addition, we showed that 36 additional proteins were causal for these risk factors. Finally, the Phe-MR highlighted additional beneficial indications of therapeutically targeting the six stroke-associated proteins and importantly, indicated few potential safety concerns. Although, as many of the phenotypes tested are not independent, the definition of significance used here might be too conservative (Bonferroni-corrected *P-*value adjusted for the number of proteins tested (six) and the total number of phenotypes (784) (*P*=0.05/6/784=1.06×10^−5^).

Tissue factor pathway inhibitor (TFPI) is primarily secreted by endothelial cells and is an anticoagulant that acts on the clotting cascade ^57^. Observational studies showed that lower levels of free TFPI were associated with higher risk of ischemic stroke ^58^ and higher risk of first and recurrent venous thrombosis ^59^, while inhibition of TFPI showed to be an effective treatment of bleeding associated with hemophilia ^60^. Consistent with this, we provided genetic evidence for directionally consistent effects of TFPI on multiple ischemic traits, such as ischemic stroke and ischemic heart disease, and opposite effects on haemorrhagic traits (*e*.*g*., gastrointestinal haemorrhage, *P*=5.23×10^−5^; excessive or frequent menstruation in females, *P*=2.70×10^−10^). We also showed that higher levels of TFPI were associated with lower BMI and WMH (**Figure 5**), and lower risk of hyperlipidemia, specifically hypercholesterolemia (**Figure 6**), suggesting that the pathways through which TFPI influences stroke risk might go beyond anticoagulation, *e*.*g*., inflammation or atherosclerotic changes. Animal studies ^60,61^ provide supporting evidence that TFPI has a role in attenuating arterial thrombus formation and atherosclerosis development. Future studies of TFPI in cardiovascular diseases focusing on the role of TFPI activity and different TFPI isoforms in the development of atherogenesis could provide further insights.

TMPRSS5 (Transmembrane Protease Serine 5, also known as Spinesin) is a member of the Type II Transmembrane Serine Protease Family (TTSPs) ^62^. For example, TMPRSS10 (Corin), one member of the TTSPs, has been reported to be involved in cardiac conduction and myometrial relaxation and contraction pathways in regulating blood pressure and promoting natriuresis, diuresis and vasodilation ^63^. Unlike Corin, the function of TMPRSS5 on cardiovascular systems is poorly understood. Human *TMPRSS5* mRNA has been shown to be expressed in the brain and the protein is predominantly expressed in neurons, in their axons in the spinal cord ^64^. A mouse model with mutant TMPRSS5 had reduced proteolytic activity and suggested a role in hearing loss ^65^. We were unable to find other studies that implicate TMPRSS5 in cardiovascular disease, both for any ischemic stroke and cardioembolic stroke, an effect that might be mediated by risk of atrial fibrillation (**Figure 4**). Furthermore, Phe-MR analysis revealed suggestive additional beneficial effects when targeted at TMPRSS5, *e*.*g*., reduced risk of Parkinson’s disease (*P*=2.15×10^−5^) and left bundle branch block (*P*=1.43×10^−5^). Taken together, TMPRSS5 represents a potentially promising therapeutic target for atrial fibrillation and cardioembolic stroke, and further research is warranted to decipher the mechanism through which it protects against cardiovascular and neurological diseases.

In addition, we have identified CD6, a lymphocyte surface receptor, associated with increased risk of any stroke. CD6 is a pan T cell marker ^66,67^, and involved in T cell proliferation and activation through its interaction with ALCAM (activated leukocyte cell adhesion molecule) ^68^. The interaction of CD6 and ALCAM is required to promote an inflammatory T cell response ^69^. Interestingly, Smedbakken L *et al* ^70^ found that acute ischemic stroke patients with upregulated ALCAM at admission had a significantly poorer survival rate (*P*<0.001). Given this interaction and that the recruitment of leukocytes and platelets is widely regarded as a pivotal step in the inflammatory response associated with cerebral ischemia ^71,72^, together with our finding that CD6 is associated with stroke, further investigation of CD6 in the context of stroke is justified.

Our study not only identified potentially novel targets (*i*.*e*. TFPI, TMPRSS5 and CD6) for stroke, but also validated proteins that had been identified as causally associated with cardiovascular diseases in previous proteome MR studies ^11,46,73^, *e*.*g*. CD40, MMP12 and IL6RA. Genetic variants in the *IL6R* region are associated with risk of inflammatory related diseases ^44^, including coronary heart disease ^74^, stroke ^73^, atrial fibrillation ^75^ and rheumatoid arthritis ^76^. Moreover, IL6R is the target of an FDA-approved therapy (Tocilizumab) for the treatment of several diseases, *e*.*g*., rheumatoid arthritis and systemic juvenile idiopathic arthritis. Phase II clinical trials testing tocilizumab for the therapy of Non-ST Elevation Myocardial Infarction have reported promising results ^77^ and a phase III clinical trial testing Ziltivekimab in cardiovascular disease and chronic kidney disease has recently started (NCT05021835).

To avoid violating the MR assumptions, we performed various sensitivity analyses. We used LD clumping at *R*^*2*^ 0.1for pQTLs with *P*5.0×10^−8^ to choose instruments for each of plasma protein level. However, concern ^78^ has been raised about the independency of the variants used as instrumental variables leading to violation of the InSIDE (instrument strength independent of direct effect) assumption of the MR-Egger method used. Therefore, we performed several sensitivity analyses to validate the robustness of the instrumental variables used in the MR analysis. Firstly, we performed MR analyses adjusting for the correlation of the variants used and obtained consistent and similar causal effect estimates to those obtained without adjusting for the correlation (**Supplementary Table 10**). Secondly, we performed conditional analysis and fine-mapping analysis to obtain instrumental variables for the six potential causal proteins and we obtained consistent MR results (**Supplementary Table 12 & Supplementary Figure 7**). Finally, colocalization analyses across the genetic associations with protein levels and stroke outcomes showed they were likely to have shared causal variants across these traits, supporting the validity of instrumental variables and the causal protein associations (**Supplementary Table 4**).

The Olink assay^19^ used in our study measures the bulk concentration of protein in plasma. However, because this assay cannot distinguish free from bound protein or active from inactive, only limited mechanistic insights can be made. Due to the limited capture of human proteome (1.5% of all known proteins), we could not evaluate the effects of all proteins within the same family or all proteins encoded within the same genomic region. For instance, we found that TMPRSS5 was a potential novel drug target for cardioembolic stroke, while other proteins in the Type II Transmembrane Serine Protease Family (TTSPs) that play crucial roles in cardiac functions ^62,79^ could not be evaluated. Thus, a targeted study of the TTSP family is warranted to comprehensively evaluate their effects in cardiovascular and neurological traits.

Our results highlight potential targets of future therapies for stroke outcomes and illustrates the relevance of proteomics in identifying drug targets. Further research is necessary to assess the viability of the six identified proteins as drug targets for stroke treatment. Additional drug targets may be uncovered as more comprehensive proteomics platforms become available and more diverse non-European ancestry populations are increasingly studied. Finally, there is an increasing need for similarly comprehensive proteomics across different tissues and organs to evaluate tissue- or organ-specific protein effects.

## Data Availability

All data produced in the present work are contained in the manuscript.

https://www.megastroke.org/index.html

https://www.leelabsg.org/resources

https://www.ebi.ac.uk/gwas/

http://diagram-consortium.org/

https://cd.hugeamp.org/

https://doi.org/10.5281/zenodo.1251813

https://genome.psych.umn.edu/index.php/GSCAN

https://app.box.com/s/1ev9iakptips70k8t4cm8j347if0ef2u

## Acknowledgements

Participants in the INTERVAL randomised controlled trial were recruited with the active collaboration of NHS Blood and Transplant England (www.nhsbt.nhs.uk), which has supported field work and other elements of the trial. DNA extraction and genotyping were co-funded by the National Institute for Health Research (NIHR), the NIHR BioResource (http://bioresource.nihr.ac.uk) and the NIHR Cambridge Biomedical Research Centre (BRC-1215-20014) [*]. The Olink® Proteomics assays were funded by Biogen, Inc. (Cambridge, MA, US). The academic coordinating centre for INTERVAL was supported by core funding from the: NIHR Blood and Transplant Research Unit in Donor Health and Genomics (NIHR BTRU-2014-10024), UK Medical Research Council (MR/L003120/1), British Heart Foundation (SP/09/002; RG/13/13/30194; RG/18/13/33946) and NIHR Cambridge BRC (BRC-1215-20014) [*] and funding from the EC-Innovative Medicines Initiative (BigData@Heart). A complete list of the investigators and contributors to the INTERVAL trial is provided in reference [**]. The academic coordinating centre would like to thank blood donor centre staff and blood donors for participating in the INTERVAL trial.

This work was supported by Health Data Research UK, which is funded by the UK Medical Research Council, Engineering and Physical Sciences Research Council, Economic and Social Research Council, Department of Health and Social Care (England), Chief Scientist Office of the Scottish Government Health and Social Care Directorates, Health and Social Care Research and Development Division (Welsh Government), Public Health Agency (Northern Ireland), British Heart Foundation and Wellcome.

*The views expressed are those of the author(s) and not necessarily those of the NIHR or the Department of Health and Social Care.

**Di Angelantonio E, Thompson SG, Kaptoge SK, Moore C, Walker M, Armitage J, Ouwehand WH, Roberts DJ, Danesh J, INTERVAL Trial Group. Efficiency and safety of varying the frequency of whole blood donation (INTERVAL): a randomised trial of 45 000 donors. Lancet. 2017 Nov 25;390(10110):2360-2371.

## Author contributions

L.C. performed the analyses and wrote the initial draft of the manuscript. J.M.M.H. designed and supervised the project. J.E.P, B.P, E.P provided data and analytical support. All authors contributed to the data preparation and critically reviewed the manuscript.

**Supplementary Figures (in separate file)**

**Supplementary Tables (in separate file)**

## References

1. Feigin, V.L. et al. Global, regional, and national burden of neurological disorders, 1990–2016: a systematic analysis for the Global Burden of Disease Study 2016. The Lancet Neurology 18, 459–480 (2019).

2. Ettehad, D. et al. Blood pressure lowering for prevention of cardiovascular disease and death: a systematic review and meta-analysis. The Lancet 387, 957–967 (2016).

3. Hankey, G.J. Stroke. The Lancet 389, 641–654 (2017).

4. Santos, R. et al. A comprehensive map of molecular drug targets. Nat Rev Drug Discov 16, 19–34 (2017).

5. Olszewski, A.J. & Szostak, W.B. Homocysteine content of plasma proteins in ischemic heart disease. Atherosclerosis 69, 109–13 (1988).

6. Robins, S.J., Lyass, A., Brocia, R.W., Massaro, J.M. & Vasan, R.S. Plasma lipid transfer proteins and cardiovascular disease. The Framingham Heart Study. Atherosclerosis 228, 230–6 (2013).

7. Goetzl, E.J. et al. Altered lysosomal proteins in neural-derived plasma exosomes in preclinical Alzheimer disease. Neurology 85, 40–7 (2015).

8. Feldreich, T. et al. Circulating proteins as predictors of cardiovascular mortality in end-stage renal disease. Journal of nephrology 32, 111–119 (2019).

9. Hauser, A.S. et al. Pharmacogenomics of GPCR Drug Targets. Cell 172, 41–54 e19 (2018).

10. Ursu, O., Glick, M. & Oprea, T. Novel drug targets in 2018. Nat Rev Drug Discov (2019).

11. Sun, B.B. et al. Genomic atlas of the human plasma proteome. Nature 558, 73–79 (2018).

12. Yao, C. et al. Genome-wide mapping of plasma protein QTLs identifies putatively causal genes and pathways for cardiovascular disease. Nat Commun 9, 3268 (2018).

13. Emilsson, V. et al. Co-regulatory networks of human serum proteins link genetics to disease. Science 361, 769–773 (2018).

14. Smith, G.D. Mendelian randomization for strengthening causal inference in observational studies: application to gene× environment interactions. Perspectives on Psychological Science 5, 527–545 (2010).

15. Davey Smith, G. & Hemani, G. Mendelian randomization: genetic anchors for causal inference in epidemiological studies. Human molecular genetics 23, R89–R98 (2014).

16. Di Angelantonio, E. et al. Efficiency and safety of varying the frequency of whole blood donation (INTERVAL): a randomised trial of 45 000 donors. The Lancet 390, 2360–2371 (2017).

17. Malik, R. et al. Multiancestry genome-wide association study of 520,000 subjects identifies 32 loci associated with stroke and stroke subtypes. Nat Genet 50, 524–537 (2018).

18. Denny, J.C. et al. Systematic comparison of phenome-wide association study of electronic medical record data and genome-wide association study data. Nat Biotechnol 31, 1102–10 (2013).

19. Assarsson, E. et al. Homogenous 96-plex PEA immunoassay exhibiting high sensitivity, specificity, and excellent scalability. PloS one 9, e95192 (2014).

20. Durbin, R. Efficient haplotype matching and storage using the positional Burrows– Wheeler transform (PBWT). Bioinformatics 30, 1266–1272 (2014).

21. Astle, W.J. et al. The allelic landscape of human blood cell trait variation and links to common complex disease. Cell 167, 1415-1429. e19 (2016).

22. Marchini, J., Howie, B., Myers, S., McVean, G. & Donnelly, P. A new multipoint method for genome-wide association studies by imputation of genotypes. Nature Genetics 39, 906–913 (2007).

23. Chang, C.C. et al. Second-generation PLINK: rising to the challenge of larger and richer datasets. Gigascience 4, 7 (2015).

24. Pierce, B.L., Ahsan, H. & Vanderweele, T.J. Power and instrument strength requirements for Mendelian randomization studies using multiple genetic variants. Int J Epidemiol 40, 740–52 (2011).

25. Palmer, T.M. et al. Using multiple genetic variants as instrumental variables for modifiable risk factors. Statistical Methods in Medical Research 21, 223–242 (2012).

26. Benner, C. et al. FINEMAP: efficient variable selection using summary data from genome-wide association studies. Bioinformatics 32, 1493–1501 (2016).

27. Surendran, P. et al. Discovery of rare variants associated with blood pressure regulation through meta-analysis of 1.3 million individuals. Nature Genetics (2020).

28. Nielsen, J.B. et al. Biobank-driven genomic discovery yields new insight into atrial fibrillation biology. Nat Genet 50, 1234–1239 (2018).

29. Mahajan, A. et al. Fine-mapping type 2 diabetes loci to single-variant resolution using high-density imputation and islet-specific epigenome maps. Nat Genet 50, 1505–1513 (2018).

30. Persyn, E. et al. Genome-wide association study of MRI markers of cerebral small vessel disease in 42,310 participants. Nature Communications 11, 2175 (2020).

31. Pulit, S.L. et al. Meta-analysis of genome-wide association studies for body fat distribution in 694 649 individuals of European ancestry. Hum Mol Genet 28, 166–174 (2019).

32. Liu, M. et al. Association studies of up to 1.2 million individuals yield new insights into the genetic etiology of tobacco and alcohol use. Nat Genet 51, 237–244 (2019).

33. Zhou, W. et al. Efficiently controlling for case-control imbalance and sample relatedness in large-scale genetic association studies. Nat Genet 50, 1335–1341 (2018).

34. Burgess, S., Butterworth, A. & Thompson, S.G. Mendelian Randomization Analysis With Multiple Genetic Variants Using Summarized Data. Genetic Epidemiology 37, 658–665 (2013).

35. Yavorska, O.O. & Burgess, S. MendelianRandomization: an R package for performing Mendelian randomization analyses using summarized data. International Journal of Epidemiology 46, 1734–1739 (2017).

36. Hemani, G. et al. The MR-Base platform supports systematic causal inference across the human phenome. Elife 7(2018).

37. Lawlor, D.A. Commentary: Two-sample Mendelian randomization: opportunities and challenges. International journal of epidemiology 45, 908 (2016).

38. Bowden, J., Smith, G.D. & Burgess, S. Mendelian randomization with invalid instruments: effect estimation and bias detection through Egger regression. International Journal of Epidemiology 44, 512–525 (2015).

39. Verbanck, M., Chen, C.-y., Neale, B. & Do, R. Detection of widespread horizontal pleiotropy in causal relationships inferred from Mendelian randomization between complex traits and diseases. Nature genetics 50, 693–698 (2018).

40. Burgess, S., Foley, C.N., Allara, E., Staley, J.R. & Howson, J.M.M. A robust and efficient method for Mendelian randomization with hundreds of genetic variants. Nature Communications 11, 376 (2020).

41. Foley, C.N. et al. A fast and efficient colocalization algorithm for identifying shared genetic risk factors across multiple traits. Nat Commun 12, 764 (2021).

42. Giambartolomei, C. et al. Bayesian test for colocalisation between pairs of genetic association studies using summary statistics. PLoS Genet 10, e1004383 (2014).

43. Zheng, J. et al. Phenome-wide Mendelian randomization mapping the influence of the plasma proteome on complex diseases. Nat Genet 52, 1122–1131 (2020).

44. Ferreira, R.C. et al. Functional IL6R 358Ala Allele Impairs Classical IL-6 Receptor Signaling and Influences Risk of Diverse Inflammatory Diseases. PLOS Genetics 9, e1003444 (2013).

45. Williams, F.M. et al. Ischemic stroke is associated with the ABO locus: the EuroCLOT study. Ann Neurol 73, 16–31 (2013).

46. Chong, M. et al. Novel Drug Targets for Ischemic Stroke Identified Through Mendelian Randomization Analysis of the Blood Proteome. Circulation (2019).

47. Hodgson, J. et al. Characterization of GDF2 Mutations and Levels of BMP9 and BMP10 in Pulmonary Arterial Hypertension. Am J Respir Crit Care Med 201, 575–585 (2020).

48. Wolf, P.A., Abbott, R.D. & Kannel, W.B. Atrial fibrillation as an independent risk factor for stroke: the Framingham Study. Stroke 22, 983–988 (1991).

49. Yang, X.-M. et al. Atrial fibrillation known before or detected after stroke share similar risk of ischemic stroke recurrence and death. Stroke 50, 1124–1129 (2019).

50. Lawes, C.M., Bennett, D.A., Feigin, V.L. & Rodgers, A. Blood pressure and stroke: an overview of published reviews. Stroke 35, 776–785 (2004).

51. Kannel, W.B., Wolf, P.A., Verter, J. & McNamara, P.M. Epidemiologic assessment of the role of blood pressure in stroke: the Framingham study. Jama 276, 1269–1278 (1996).

52. Mäntylä, R. et al. Magnetic resonance imaging white matter hyperintensities and mechanism of ischemic stroke. Stroke 30, 2053–2058 (1999).

53. Mitchell, A.B. et al. Obesity Increases Risk of Ischemic Stroke in Young Adults. Stroke 46, 1690–1692 (2015).

54. Kivimäki, M. et al. Overweight, obesity, and risk of cardiometabolic multimorbidity: pooled analysis of individual-level data for 120 813 adults from 16 cohort studies from the USA and Europe. The Lancet Public Health 2, e277–e285 (2017).

55. Janghorbani, M. et al. Prospective Study of Type 1 and Type 2 Diabetes and Risk of Stroke Subtypes. The Nurses’ Health Study 30, 1730–1735 (2007).

56. Rost, N.S. et al. White matter hyperintensity volume is increased in small vessel stroke subtypes. Neurology 75, 1670–1677 (2010).

57. Broze Jr, G.J. Tissue factor pathway inhibitor. Thrombosis and haemostasis 73, 090–093 (1995).

58. He, M. et al. Observation on tissue factor pathway and some other coagulation parameters during the onset of acute cerebrocardiac thrombotic diseases. Thrombosis research 107, 223–228 (2002).

59. Hoke, M. et al. Tissue factor pathway inhibitor and the risk of recurrent venous thromboembolism. Thrombosis and haemostasis 94, 787–790 (2005).

60. Waters, E.K. et al. Aptamer ARC19499 mediates a procoagulant hemostatic effect by inhibiting tissue factor pathway inhibitor. Blood 117, 5514–5522 (2011).

61. Westrick, R.J. et al. Deficiency of tissue factor pathway inhibitor promotes atherosclerosis and thrombosis in mice. Circulation 103, 3044–3046 (2001).

62. Bugge, T.H., Antalis, T.M. & Wu, Q. Type II transmembrane serine proteases. J Biol Chem 284, 23177–81 (2009).

63. Knappe, S., Wu, F., Masikat, M.R., Morser, J. & Wu, Q. Functional analysis of the transmembrane domain and activation cleavage of human corin design and characterization of a soluble corin. Journal of Biological Chemistry 278, 52363–52370 (2003).

64. Yamaguchi, N., Okui, A., Yamada, T., Nakazato, H. & Mitsui, S. Spinesin/TMPRSS5, a novel transmembrane serine protease, cloned from human spinal cord. J Biol Chem 277, 6806–12 (2002).

65. Guipponi, M. et al. An integrated genetic and functional analysis of the role of type II transmembrane serine proteases (TMPRSSs) in hearing loss. Hum Mutat 29, 130–41 (2008).

66. Carrasco, E. et al. Human CD6 Down-Modulation following T-Cell Activation Compromises Lymphocyte Survival and Proliferative Responses. Front Immunol 8, 769 (2017).

67. Hernández, P., Moreno, E., Aira, L.E. & Rodríguez, P.C. Therapeutic Targeting of CD6 in Autoimmune Diseases: A Review of Cuban Clinical Studies with the Antibodies IOR-T1 and Itolizumab. Curr Drug Targets 17, 666–77 (2016).

68. Zimmerman, A.W. et al. Long-term engagement of CD6 and ALCAM is essential for T-cell proliferation induced by dendritic cells. Blood 107, 3212–3220 (2006).

69. Gimferrer, I. et al. Relevance of CD6-Mediated Interactions in T Cell Activation and Proliferation. The Journal of Immunology 173, 2262–2270 (2004).

70. Smedbakken, L. et al. Activated Leukocyte Cell Adhesion Molecule and Prognosis in Acute Ischemic Stroke. Stroke 42, 2453–2458 (2011).

71. Jin, R., Yang, G. & Li, G. Inflammatory mechanisms in ischemic stroke: role of inflammatory cells. J Leukoc Biol 87, 779–89 (2010).

72. Elkind, M.S. Inflammatory mechanisms of stroke. Stroke 41, S3–8 (2010).

73. Georgakis, M.K. et al. Genetically determined levels of circulating cytokines and risk of stroke: role of monocyte chemoattractant protein-1. Circulation 139, 256–268 (2019).

74. Swerdlow, D.I. et al. The interleukin-6 receptor as a target for prevention of coronary heart disease: a mendelian randomisation analysis. Lancet 379, 1214–24 (2012).

75. Schnabel, R.B. et al. Large-scale candidate gene analysis in whites and African Americans identifies IL6R polymorphism in relation to atrial fibrillation: the National Heart, Lung, and Blood Institute’s Candidate Gene Association Resource (CARe) project. Circulation: Cardiovascular Genetics 4, 557–564 (2011).

76. Eyre, S. et al. High-density genetic mapping identifies new susceptibility loci for rheumatoid arthritis. Nature genetics 44, 1336–1340 (2012).

77. Ueland, T. et al. Serum PCSK9 is modified by interleukin-6 receptor antagonism in patients with hypercholesterolaemia following non-ST-elevation myocardial infarction. Open heart 5, e000765 (2018).

78. Plump, A. & Davey Smith, G. Identifying and Validating New Drug Targets for Stroke and Beyond: Can Mendelian Randomization Help? (Am Heart Assoc, 2019).

79. Szabo, R. et al. Type II transmembrane serine proteases. Thromb Haemost 90, 185–93 (2003).

