## Supplemental Figures for "Genetically predicted levels of the human plasma proteome and risk of stroke: a Mendelian Randomization study"

### Supplementary Figures

#### Supplementary Figure 1

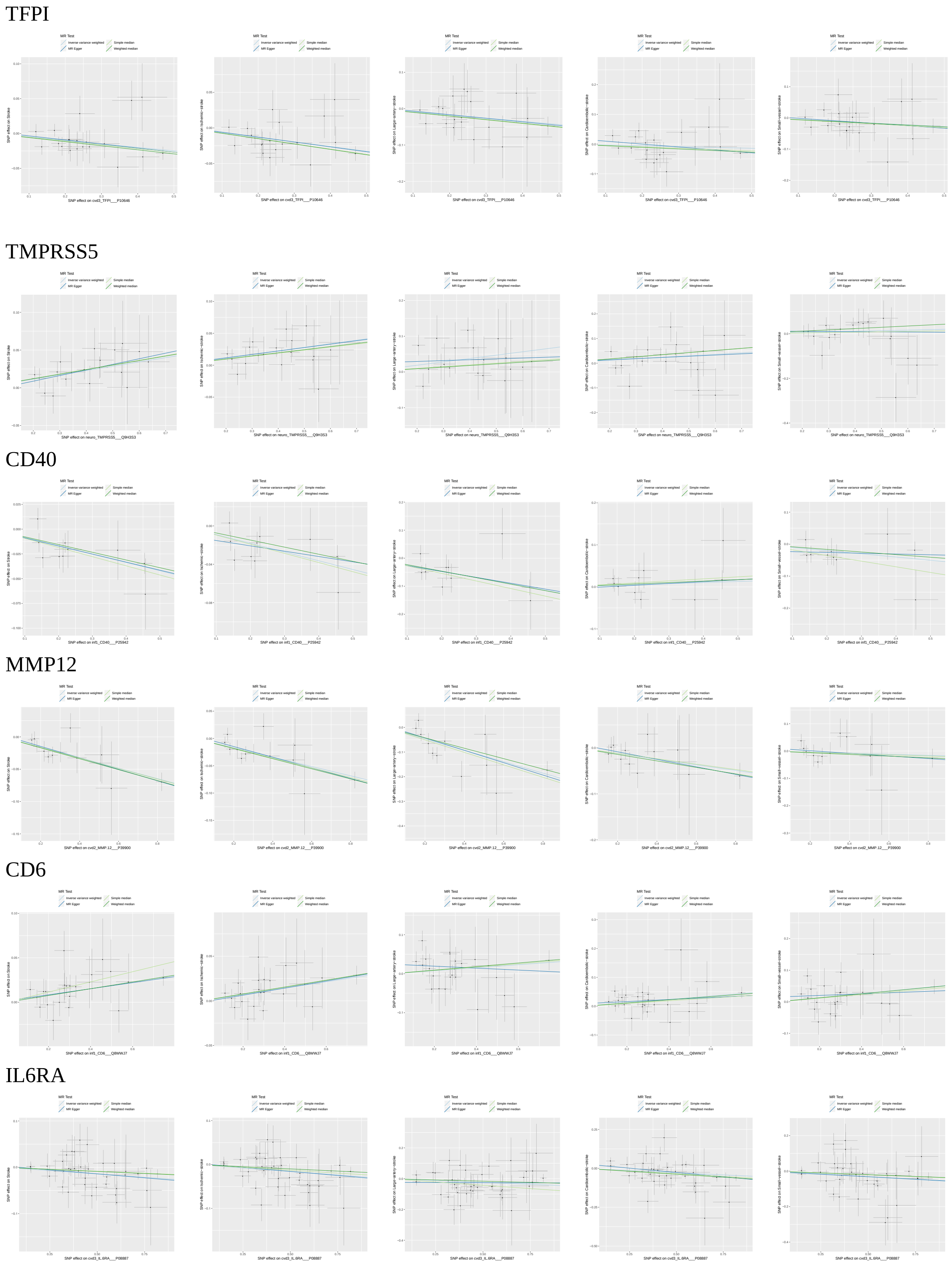


**Supplementary Figure 1.** Standard MR plots for primary MR results: the causal effect of proteins on stroke outcomes.

**Supplementary Figure 2**

A

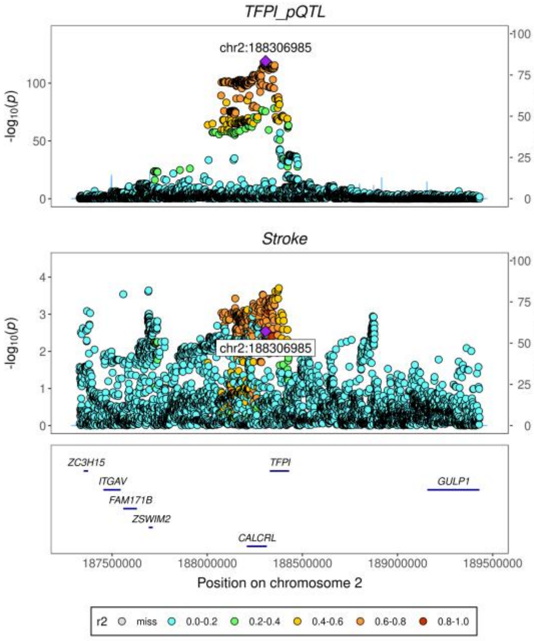


C

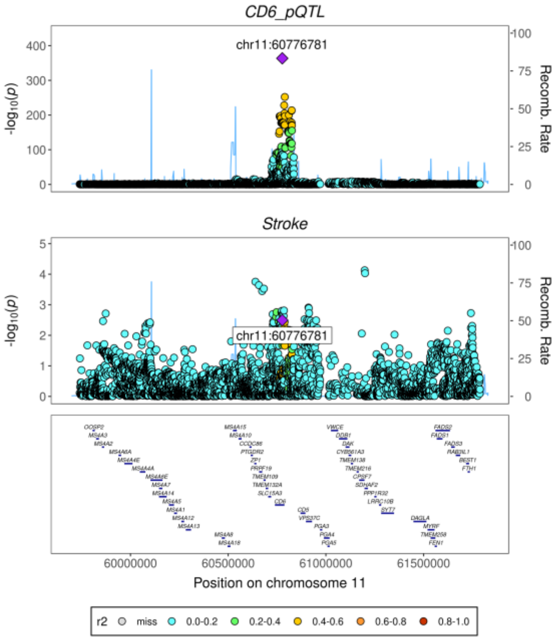


B

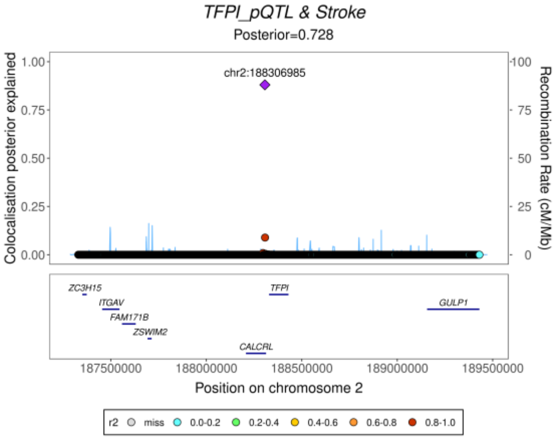


D

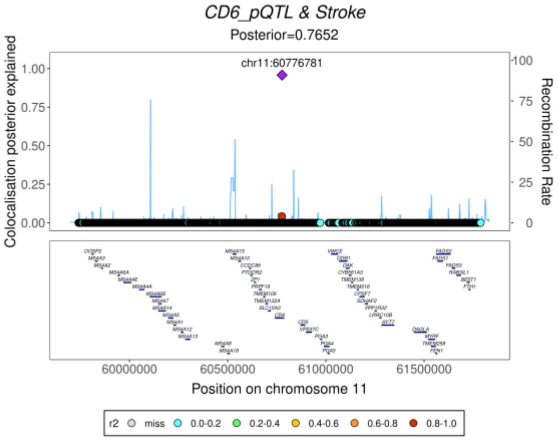

E

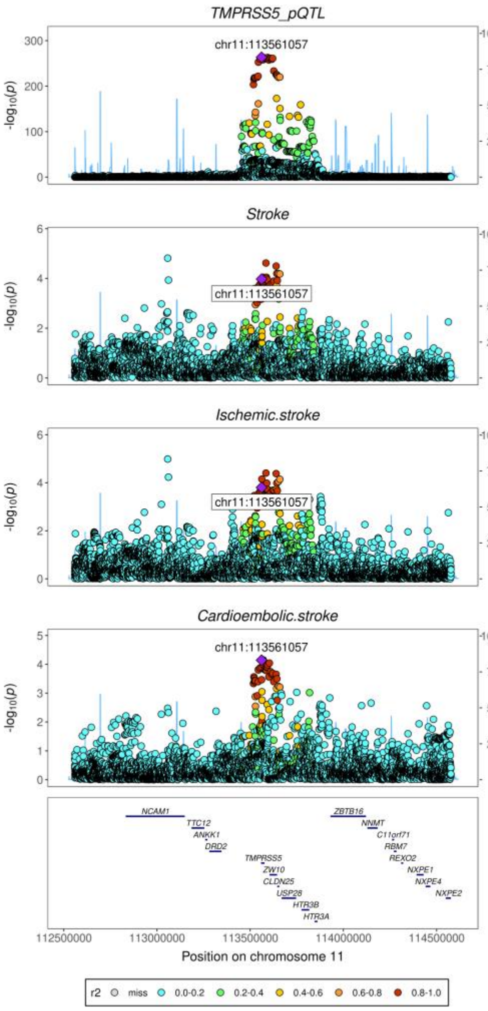
TMPRSS5_pQTL
chr11:113561057
-log10(p)
Recomb. Rate
Stroke
chr11:113561057
Ischemic.stroke
chr11:113561057
Cardioembolic.stroke
chr11:113561057
NCAM1
TTC12
ANKK1
DRD2
TMPRSS5
ZW10
CLDN25
USP28
HTR3B
HTR3A
ZBTB16
NNMT
C11orf71
RBM7
REXO2
NXPE1
NXPE4
NXPE2
112500000 113000000 113500000 114000000 114500000
Position on chromosome 11
r2 miss 0.0-0.2 0.2-0.4 0.4-0.6 0.6-0.8 0.8-1.0

G

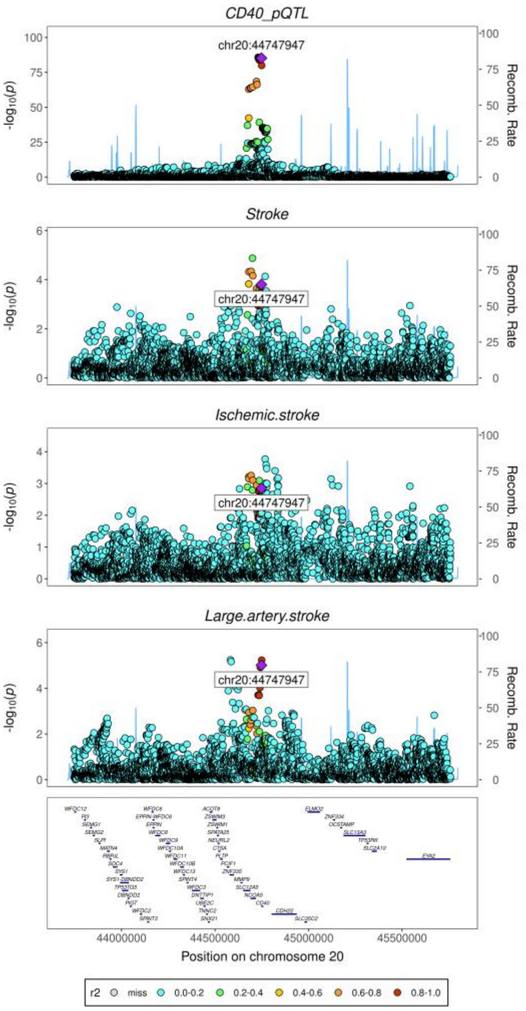
CD40_pQTL
chr20:44747947
-log10(p)
Recomb. Rate
Stroke
chr20:44747947
Ischemic.stroke
chr20:44747947
Large.artery.stroke
chr20:44747947
44000000 44500000 45000000 45500000
Position on chromosome 20
r2 miss 0.0-0.2 0.2-0.4 0.4-0.6 0.6-0.8 0.8-1.0

F

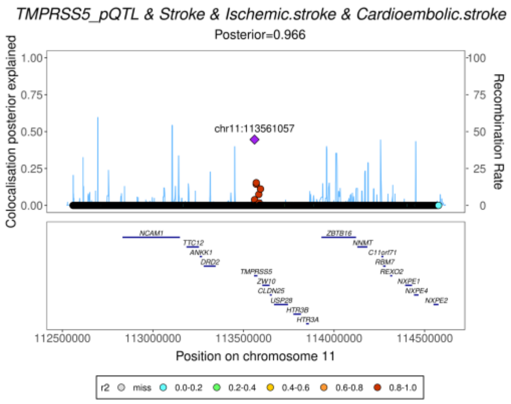
TMPRSS5_pQTL & Stroke & Ischemic.stroke & Cardioembolic.stroke
Posterior=0.966
Colocalisation posterior explained
Recombination Rate
chr11:113561057
NCAM1
TTC12
ANKK1
DRD2
TMPRSS5
ZW10
CLDN25
USP28
HTR3B
HTR3A
ZBTB16
NNMT
C11orf71
RBM7
REXO2
NXPE1
NXPE4
NXPE2
112500000 113000000 113500000 114000000 114500000
Position on chromosome 11
r2 miss 0.0-0.2 0.2-0.4 0.4-0.6 0.6-0.8 0.8-1.0

H

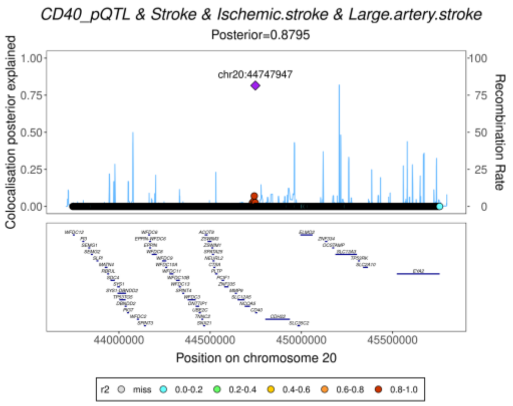
CD40_pQTL & Stroke & Ischemic.stroke & Large.artery.stroke
Posterior=0.8795
Colocalisation posterior explained
Recombination Rate
chr20:44747947
44000000 44500000 45000000 45500000
Position on chromosome 20
r2 miss 0.0-0.2 0.2-0.4 0.4-0.6 0.6-0.8 0.8-1.0

I

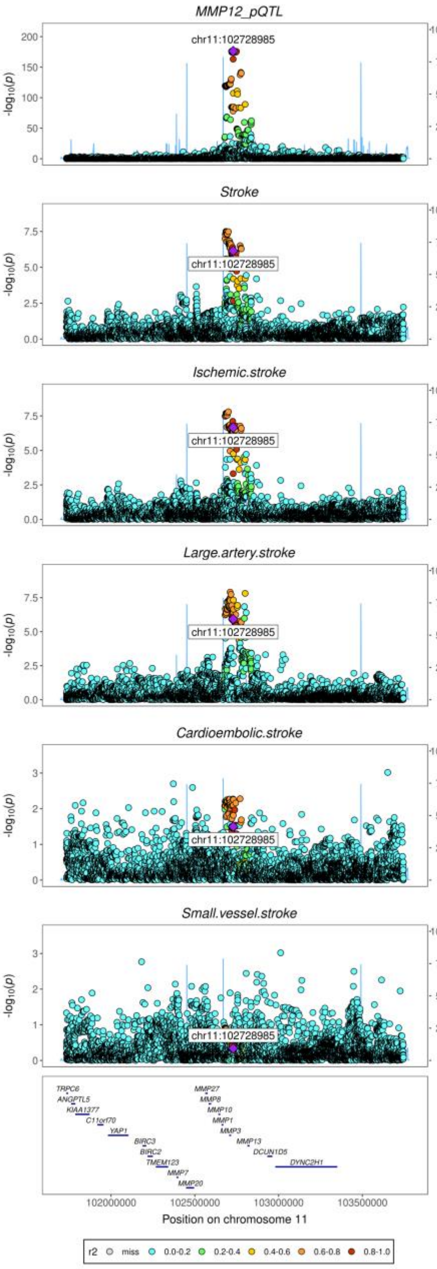
MMP12_pQTL
Stroke
Ischemic.stroke
Large.artery.stroke
Cardioembolic.stroke
Small.vessel.stroke
chr11:102728985
-log10(p)
Recomb. Rate
TRPC6
ANGPTL5
KIAA1377
C11orf70
YAP1
BIRC3
BIRC2
TMEM123
MMP7
MMP20
MMP27
MMP8
MMP10
MMP1
MMP3
MMP13
DCUN1D5
DYNC2H1
102000000
102500000
103000000
103500000
Position on chromosome 11
r2 miss 0.0-0.2 0.2-0.4 0.4-0.6 0.6-0.8 0.8-1.0

K

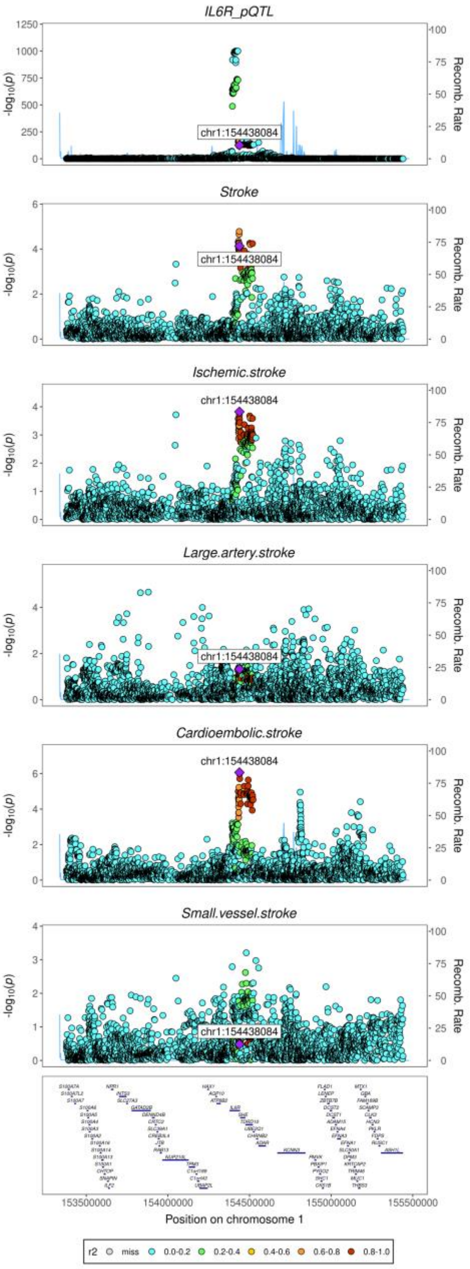
IL6R_pQTL
Stroke
Ischemic.stroke
Large.artery.stroke
Cardioembolic.stroke
Small.vessel.stroke
chr1:154438084
-log10(p)
Recomb. Rate
153500000
154000000
154500000
155000000
155500000
Position on chromosome 1
r2 miss 0.0-0.2 0.2-0.4 0.4-0.6 0.6-0.8 0.8-1.0

J

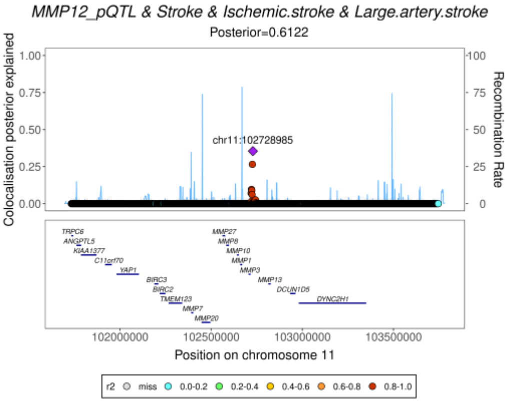
MMP12_pQTL & Stroke & Ischemic.stroke & Large.artery.stroke
Posterior=0.6122
chr11:102728985
Colocalisation posterior explained
Recombination Rate
TRPC6
ANGPTL5
KIAA1377
C11orf70
YAP1
BIRC3
BIRC2
TMEM123
MMP7
MMP20
MMP27
MMP8
MMP10
MMP1
MMP3
MMP13
DCUN1D5
DYNC2H1
102000000
102500000
103000000
103500000
Position on chromosome 11
r2 miss 0.0-0.2 0.2-0.4 0.4-0.6 0.6-0.8 0.8-1.0

L

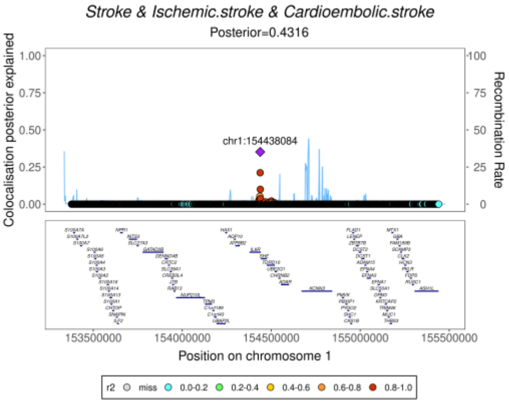
Stroke & Ischemic.stroke & Cardioembolic.stroke
Posterior=0.4316
chr1:154438084
Colocalisation posterior explained
Recombination Rate
153500000
154000000
154500000
155000000
155500000
Position on chromosome 1
r2 miss 0.0-0.2 0.2-0.4 0.4-0.6 0.6-0.8 0.8-1.0

**Supplementary Figure 2**. Colocalization plots of pQTLs and stroke association signals.
A & B. Colocalization of TFPI pQTLs and Stroke association signals
C & D. Colocalization of CD6 pQTLs and Stroke association signals.
E & F. Colocalization of TMPRSS5 pQTLs and Stroke association signals.
G & H. Colocalization of CD40 pQTLs and Stroke association signals.
I & J. Colocalization of MMP12 pQTLs and Stroke association signals.
K & L. Colocalization of IL6RA pQTLs and Stroke association signals.
A/C/E/G/I/K: Regional association plot of pQTLs and stroke association. SNPs are plotted by their positions on the chromosome against association with the protein level, and Stroke ($-\log_{10}$ P value) on the left y axis. Points are coloured by their local linkage disequilibrium (LD) pattern with their "candidate causal SNP" (purple diamond). Blow the main plot is the track showing the position and orientation of local genes.
B/D/F/H/J/L: Plot of percentage of posterior probability (PP) of colocalization explained by each of the SNP within the corresponding region. The traits that colocalized are presented in the title of the plot and the PP of colocalization is presented in the subtitle of the plot. SNPs are plotted by their positions on the chromosome against the percentage explained on the left y axis. The best candidate causal SNP is presented in purple diamond.

**Supplementary Figure 3**

SBP

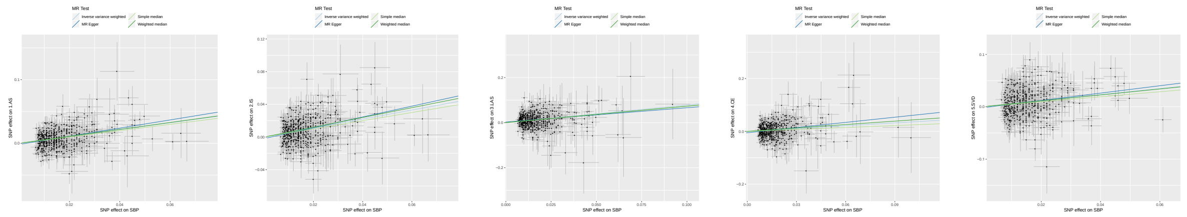

AF

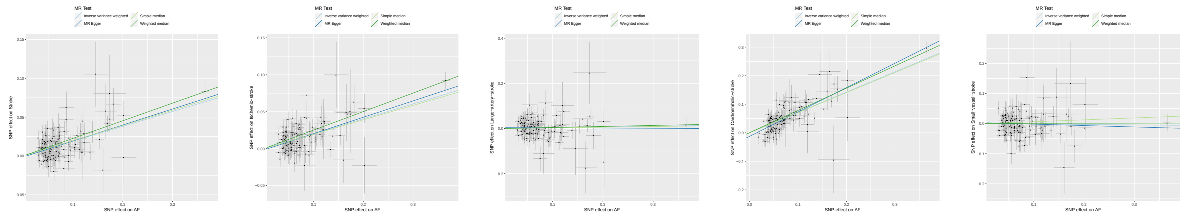

T2D

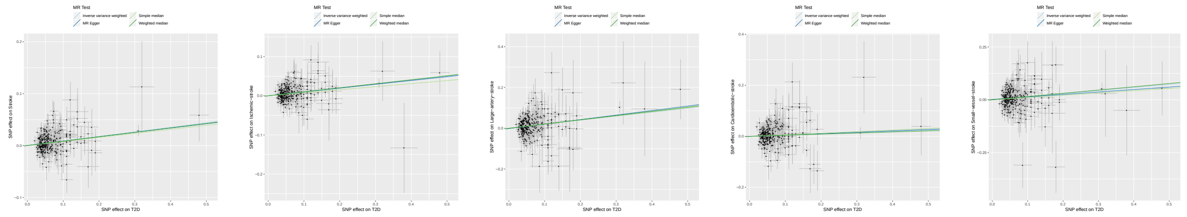

BMI

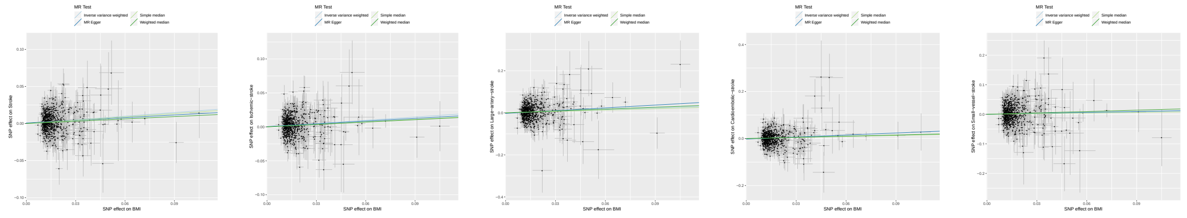

WMH

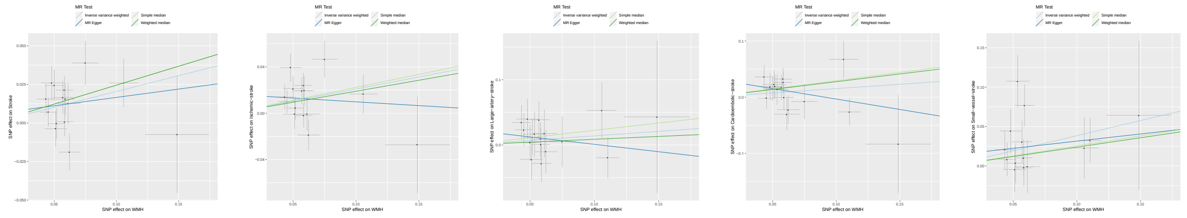

SmokingInitiation

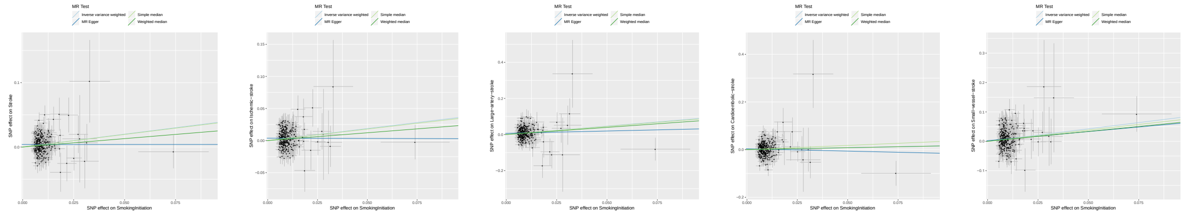

**Supplementary Figure 3.** Standard MR plots for MR results: the causal effect of risk factors on stroke outcomes.

#### Supplementary Figure 4

TFPI and BMI

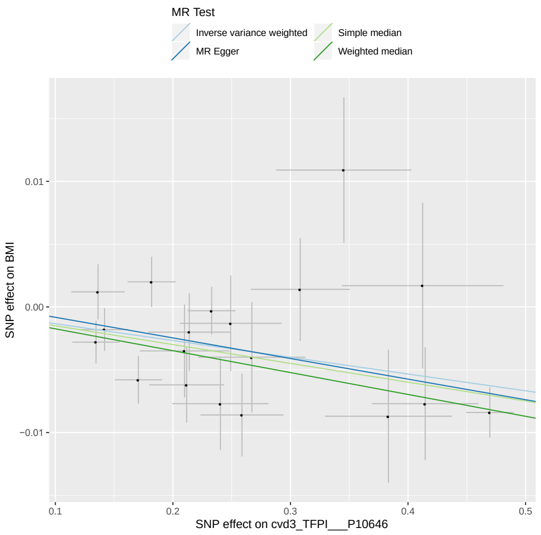


TFPI and WMH

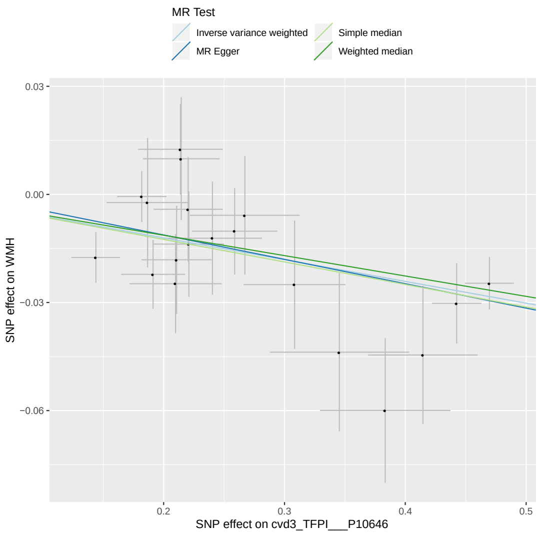


TMPRSS5 and AF

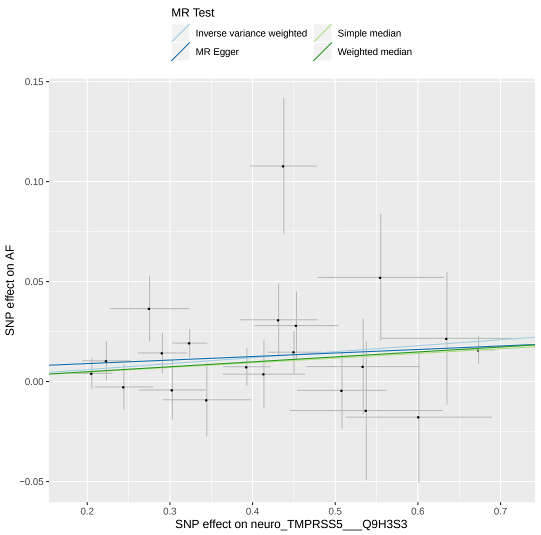


IL6RA and AF

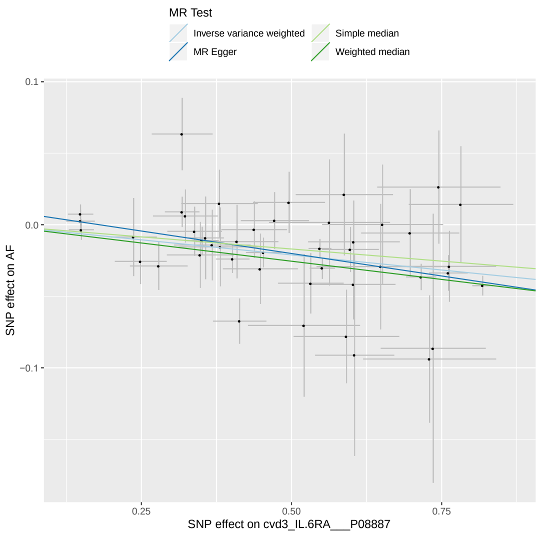


**Supplementary Figure 4.** Standard MR plots for MR results: the causal effect of proteins (stroke associated) on risk factors.

### Supplementary Figure 5

#### TFPI

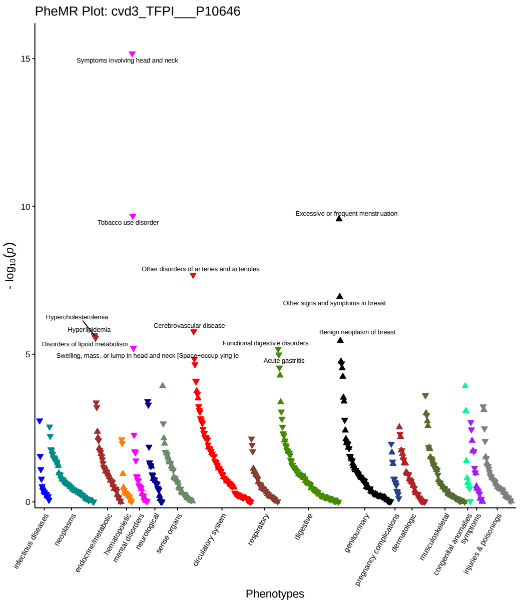


#### TMPRSS5

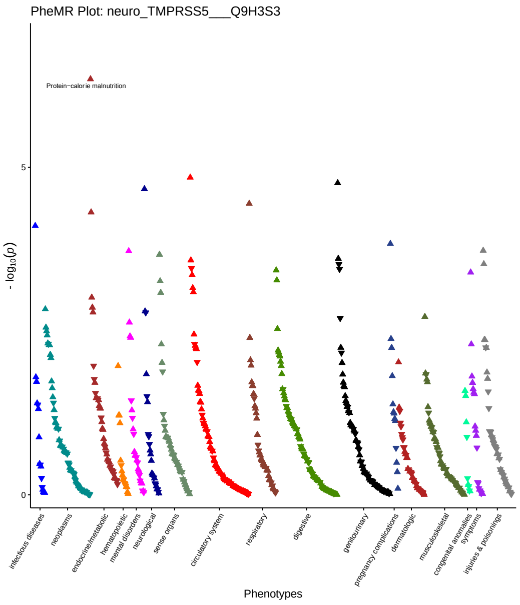


#### MMP12

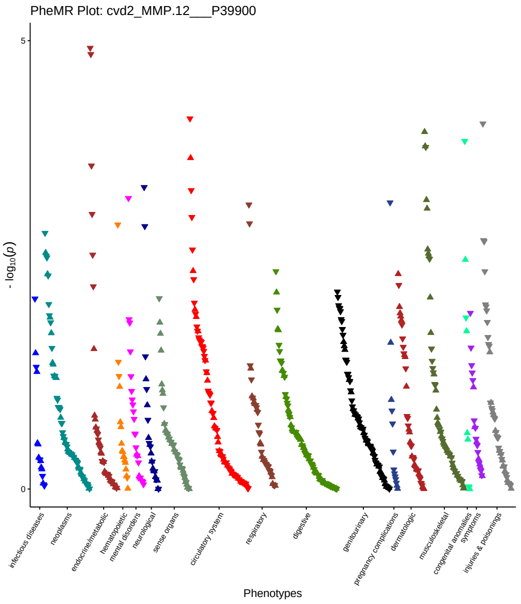


## CD40

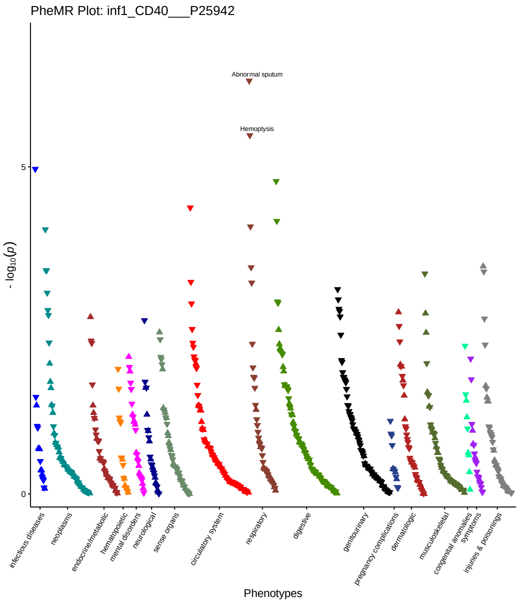

IL6RA

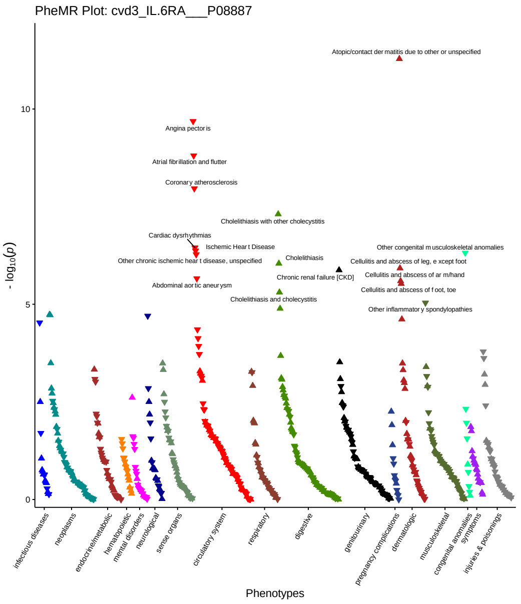


CD6

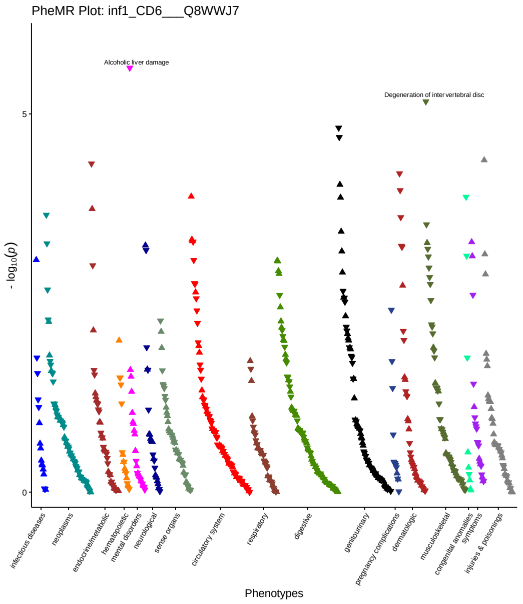


**Supplementary Figure 5**. Manhattan plots of the potential on-target side-effects from Phe-MR analysis.

Results can be perceived as the effects of per SD higher circulating protein level on each phenotype - triangle up represent risk-conferring effects and triangle down represents protective effects. If the effect direction of the target protein on the phenotype is consistent with that on stroke outcomes, it represents “beneficial” additional indications through intervention of circulating protein level. Conversely, opposing effect directions of the target protein on the phenotype and stroke represents “deleterious” side-effects. For example, a higher level of TFPI is associated with lower risk of ischemic stroke and so downface triangle represents “beneficial effects”, while the upward triangle represents “deleterious effects” when the hypothetical intervention increases TFPI levels. Associations above the pink line passed Bonferroni significance $P \leq 0.05/6/784 = 1.06\times10^{-5}$).

#### Supplementary Figure 6

### CD40

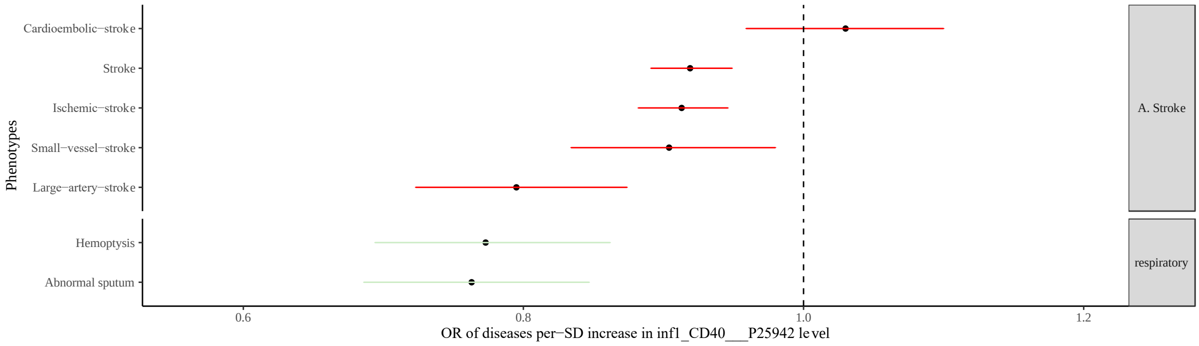


### CD6

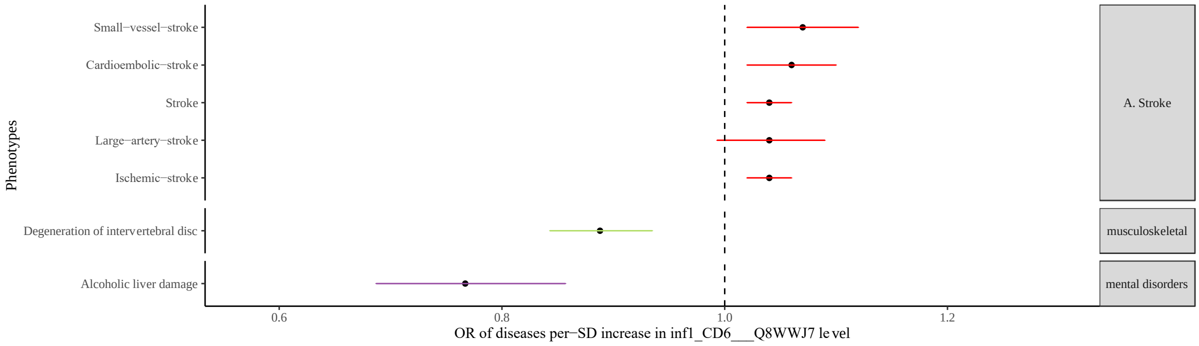


##### MMP12

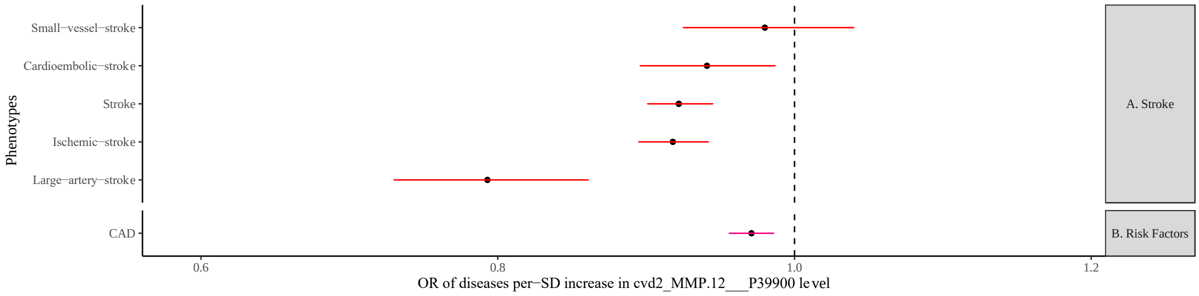

**IL6RA**

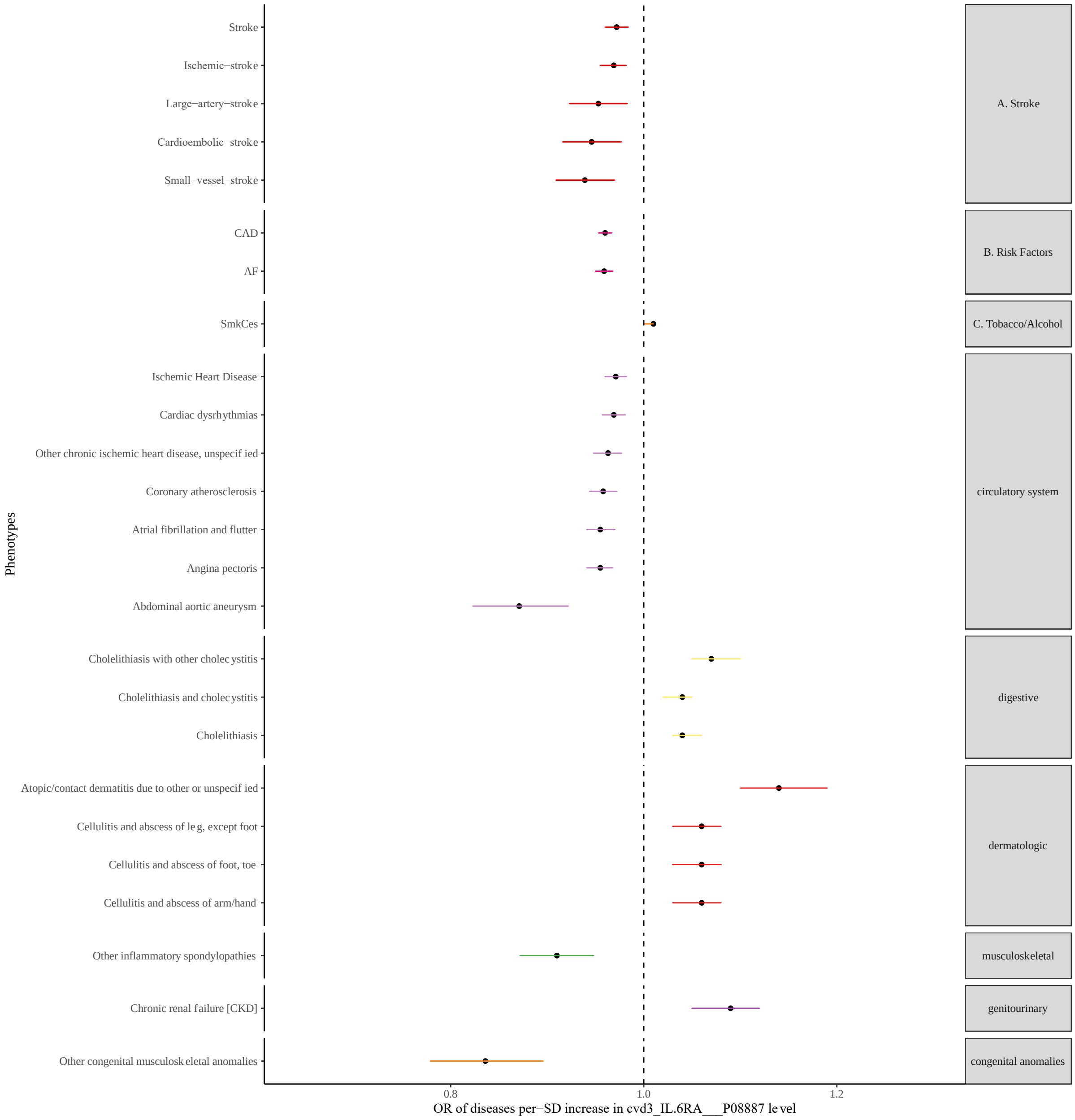


**Supplementary Figure 6**. Forest plots illustrating the potential on-target side-effects associated with causal proteins revealed by Phe-MR analysis.

Results can be perceived as the effects of per SD higher circulating protein level on each phenotype. If the effect direction of the target protein on the phenotype is consistent with that on stroke outcomes, it represents “beneficial” additional indications through intervention of circulating protein level. Conversely, opposing effect directions of the target protein on the phenotype and stroke represents “deleterious” side-effects. For example, a higher level of TFPI is associated with lower risk of ischemic stroke and so phenotypes with OR<1 represents “beneficial effects”, OR>1 represents “deleterious effects” when the hypothetical intervention increases TFPI levels. Only significant associations that passed Bonferroni correction ($P \leq 0.05/6/784 = 1.06\times10^{-5}$) were plotted.

#### Supplementary Figure 7

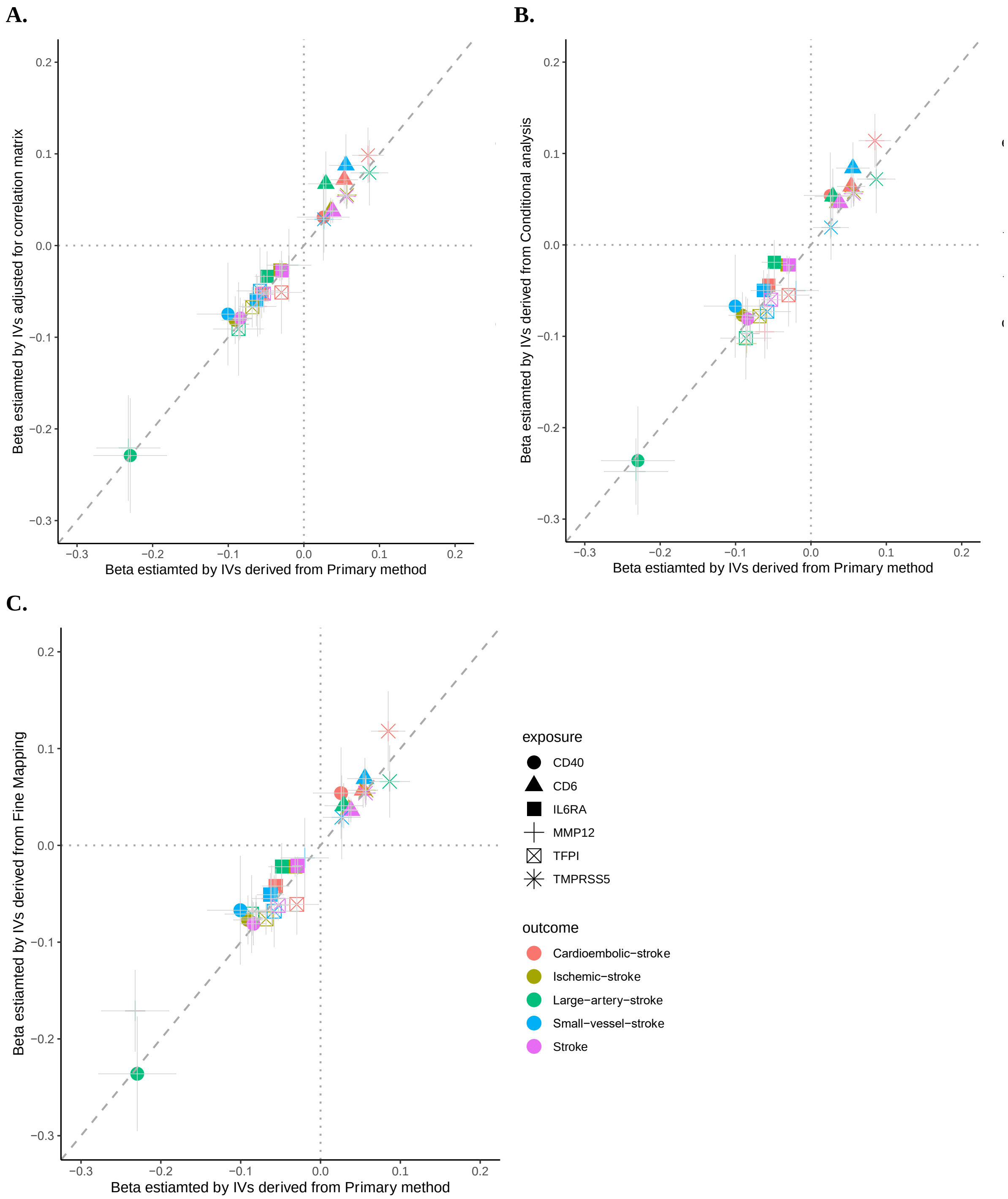


**Supplementary Figure 7**. Correlations of causal effect sizes of six proteins on stroke outcomes with IVs derived from primary method and others.

Primary method (IVs derived from LD clumping at pQTLs < $5\times10^{-8}$ at $R^2 < 0.1$) versus IVs adjusted for correlation matrix (**A**); IVs derived from conditional analysis (**B**); and IVs derived from fine mapping (**C**). Scatter plots show the effect size (plotted as beta and se) of the proteins on stroke outcomes, with each shape represents each protein and each colour represents each stroke outcome. The dashed grey line is a reference line representing y=x.
